# Genetic heterogeneity and subtypes of major depression

**DOI:** 10.1101/2021.03.05.21252911

**Authors:** Thuy-Dung Nguyen, Arvid Harder, Ying Xiong, Kaarina Kowalec, Sara Hägg, Na Cai, Ralf Kuja-Halkola, Christina Dalman, Patrick F Sullivan, Yi Lu

**Author notes:** **Corresponding author:** Yi Lu, Ph.D., Nobels Väg 12A, Solna, Sweden 17177.

## Abstract

Major depression (MD) is a heterogeneous disorder; however, the extent to which genetic factors distinguish MD patient subgroups (genetic heterogeneity) remains uncertain. This study sought evidence for genetic heterogeneity in MD. Using UK Biobank cohort, the authors defined 16 MD subtypes within eight comparison groups (vegetative symptoms, symptom severity, comorbid anxiety disorder, age at onset, recurrence, suicidality, impairment and postpartum depression; N∼3 000-47 000). To compare genetic component of these subtypes, subtype-specific genome-wide association studies were performed to estimate SNP-heritability, and genetic correlations within subtype comparison and with other related disorders or traits. The findings indicated that MD subtypes were divergent in their SNP-heritability, and genetic correlations both within subtype comparisons and with other related disorders/traits. Three subtype comparisons (vegetative symptoms, age at onset, and impairment) showed significant differences in SNP-heritability; while genetic correlations within subtype comparisons ranged from 0.55 to 0.86, suggesting genetic profiles are only partially shared among MD subtypes. Furthermore, subtypes that are more clinically challenging, e.g., early-onset, recurrent, suicidal, more severely impaired, had stronger genetic correlations with other psychiatric disorders. MD with atypical-like features showed a positive genetic correlation (*+*0.40) with BMI while a negative correlation (−0.09) was found in those without atypical-like features. Novel genomic loci with subtype-specific effects were identified. These results provide the most comprehensive evidence to date for genetic heterogeneity within MD, and suggest that the phenotypic complexity of MD can be effectively reduced by studying the subtypes which share partially distinct etiologies.

## INTRODUCTION

Major depression (MD) is a common psychiatric disorder that affects 15% of the population during lifetime.^1^ Individuals with MD vary considerably in symptoms, severity, course, treatment response, and neurobiology.^2^ MD heterogeneity is a major research and clinical challenge.^3^ Despite major efforts in epidemiological, clinical, and biological psychiatry, this decades-long challenge remains largely unresolved.^4-6^ MD subtypes have been proposed within five major categories that focused on: symptoms (typical versus atypical which characterized by improved mood in response to positive events, weight gain, increased appetite, and hypersomnia; with or without concomitant anxiety, etc.), etiology (with or without trauma or postpartum exposure), time of onset/time course (early-versus late-onset, recurrent), sex, and treatment outcome (treatment responsive versus resistant).^6^ Many of these subtypes, however, exhibit unclear distinctions in underlying biology, psychosocial factors, and treatment efficacy.^6^ One of the key biological component is genetics—the extent to which genetic factors distinguish these MD subtypes (i.e. genetic heterogeneity) is largely unknown. Given its relatively low heritability (30-40%)^7, 8^, identifying MD subtypes that are more heritable is of particular importance. Among the proposed subtypes, the sex difference in heritability is the most intensively studied, and current findings support that MD is more heritable in women than in men.^9^ Early-onset, recurrent MD, and postpartum depression have been suggested to confer higher genetic liability from family-based studies, which was subsequently confirmed using polygenic risk scores (PRS) in recent MD genome-wide association studies (GWAS).^9-13^ Comparisons of MD subtypes between early-versus late-onset, atypical versus non-atypical, with or without adversity have yielded interesting findings (e.g., the genetic overlap with metabolic traits was only found in MD with atypical features subtype, but not among those with non-atypical symptoms).^14^ The studies to-date that have used genetic approaches to index the heterogeneity of MD subtypes are encouraging (summarized in *Supplementary Table S1*) but overall impeded by a paucity of large cohorts with similar ascertainment, phenotyping, and genotyping.^5^ As a result, a systematic comparison across the MD subtypes is lacking and overall evidence for genetic heterogeneity within MD is inconclusive.

The goal of this study was to investigate genetic heterogeneity in clinically-informed MD subtypes. To accomplish this, we systematically evaluated 16 subtypes in the unique UK Biobank (UKB) cohort with large-scale genomic data and a wide array of phenotypic measures uniformly assessed. In particular, we compared genetic components among subtypes by quantifying differences in heritability (*i*.*e*., measuring the relative importance of genetic effects on phenotypic variance) and estimating genetic correlations (*i*.*e*., to determine if underlying genetic risk factors are identical) within subtype comparisons and with other traits.

## MATERIALS and METHODS

To identify MD subtypes and compare their genetic components, we carefully selected phenotypes and large-scale genotype data from the UKB. The full protocol and scripts are available via Github.

### Participant and phenotype definitions

UKB is a population-based cohort of over 500 000 adults (age 37-73) from across the United Kingdom.^15^ UKB has phenotypic data from questionnaires, health records, biological sampling, and physical measurements. Information about mental health including MD was collected using various sources, including touchscreen questionnaires, nurse interviews, hospital admission records, and web-based mental health questionnaires (MHQ) follow-up. The UKB data profile were available elsewhere^15^ and briefly described in *Supplementary Methods S1*.*1*.

#### MD case definition

Cases were identified using five MD definitions, including (i) lifetime MD based on the Composite International Diagnostic Interview (CIDI) Short Form; (ii) ICD-coded MD based on linked hospital admission records; (iii) Probable MD based on Smith et al.^16^; (iv) Self-reported MD as part of past and current medical conditions; and (v) MD cardinal symptoms of anhedonia and dysphoria (*Supplementary Table S2)*. These MD definitions have been used in previous studies.^17-19^ Because some definitions were available only for parts of the UKB samples, to maximize sample size for MD subtypes, we included individuals who met criteria for at least one of the five MD definitions as cases. MD subtypes were all nested in the broad MD group but coming from different MD definitions (*Supplementary Table S3)*.

#### MD subtypes

According to major clinical features in MD, we defined 16 MD subtypes within eight comparison dimensions including (i) MD with versus without atypical-like features based on vegetative symptoms of hypersomnia and weight gain; (ii) severe versus mild/moderate MD based on symptom severity defined in Smith et al.^16^ or ICD codes; (iii) MD with or without comorbid anxiety disorder either self-reported or based on ICD codes; (iv) early- (≤30 years old) versus late-onset (≥44 years old) MD based on age at which first experienced a ≥2-week episode of cardinal symptoms; (v) recurrent MD vs single-episode MD based on the number of episodes self-reported or ICD codes; (vi) MD with or without suicidal thoughts or self-harm either experienced recently or during the worst episode; (vii) MD with mild, moderate, severe impairment on normal roles; and (viii) postpartum depression (PPD), either self-reported or based on ICD codes (***Table 1***; *Supplementary Methods S1*.*1, Table S4*). The majority of these subtypes are included in the five major categories proposed in the previous meta-review; while the subtypes on suicidality and on impairment are considered as outcome-based subtypes (*Supplementary Table S1)*.^6^

**Table 1.** MD subtypes and sample sizes.

| Subtype | <sup>†</sup> Definition | N<sub>case</sub> | <sup>†</sup> N<sub>control</sub> |
| --- | --- | --- | --- |
| **Vegetative symptoms** |  |  |  |
| Atypical-like features | MD cases who reported both hypersomnia and weight gain | 2904 | 250229 |
| Non-atypical-like features | MD cases who did not report both hypersomnia and weight gain | 46900 | 250229 |
| **Symptom severity** |  |  |  |
| Severe | Probable recurrent MD (severe) defined by Smith et al.<sup>16</sup>; and/or ICD-diagnoses of severe MD (F322, F323, F332, F333) | 7923 | 250229 |
| Mild/moderate | Probable recurrent MD (moderate) defined by Smith et al.<sup>16</sup>, and/or ICD-diagnoses of mild (F320, F330) or moderate depression episode (F321, F331) | 11300 | 250229 |
| **Comorbid anxiety disorder** |  |  |  |
| MD with comorbid anxiety, panic attacks, phobia | MD cases with reported social anxiety/phobia, panic attacks, and anxiety, nerves/generalized anxiety disorder, and/or ICD diagnoses of anxiety disorder (F40, F41) | 24543 | 249062 |
| MD without comorbid anxiety, panic attacks, phobia | MD cases with neither self-reported nor ICD-codes anxiety disorder | 16480 | 249062 |
| **Age at onset** |  |  |  |
| Early onset ≤ 30 years old | First 3 octiles of age at which first experiencing a ≥2-week episode of cardinal symptoms | 29292 | 250229 |
| Late onset ≥ 44 years old | Last 3 octiles of age at which first experiencing a ≥2-week episode of cardinal symptoms | 27796 | 250229 |
| **Recurrence** |  |  |  |
| Recurrent episode MD | Probable MD cases with recurrent episodes, and/or with ≥2 episodes of at least two weeks of cardinal symptoms, and/or ICD-diagnosis of recurrent MD (F33) | 30219 | 250229 |
| Single episode MD | MD cases with one episode of feeling depressed, and/or self-reported a single episode of cardinal symptoms, and/or ICD-diagnosis of non-recurrent MD (F32) | 20973 | 250229 |
| **Suicidality** |  |  |  |
| MD with suicidal thoughts | MD cases with reported thoughts of death during worst depression; and/or those with recent thoughts of suicide or self-harm | 40976 | 250229 |
| MD without suicidal thoughts | MD cases without suicidal thoughts as defined above | 37140 | 250229 |
| **Impairment** |  |  |  |
| Mild impairment | Impact of MD on normal roles, including study/employment, childcare and housework, leisure pursuits, during the worst period of depression as 'not at all/a little impact' | 28721 | 250229 |
| Moderate impairment | Impact of MD is 'somewhat' | 28991 | 250229 |
| Severe impairment | Impact of MD is 'a lot' | 25825 | 250229 |
| **Postpartum** |  |  |  |
| MD related to childbirth | Women who reported post-natal depression during the nurse interview at the baseline recruitment; and/or MD cardinal symptoms related to childbirth; and/or had ICD diagnosis of mental and behavioral disorders associated with the puerperium. | 6333 | 95736 |

<sup>†</sup> Method details for deriving subtypes available in *Supplementary Table S4* and control groups in *Supplementary Table S2*

#### Control group

We used a common control group without lifetime history of MD to compare with all but the subtypes of comorbid anxiety disorder and PPD. From the entire UKB population, we excluded those with any indications of MD using five MD case criteria described above, and two additional exclusion criteria, help-seeking MD and antidepressant use (medication list in *Supplementary Table S5*). We further excluded those with ICD-diagnoses of anxiety disorders from the controls for the MD subtype with or without comorbid anxiety disorder. For PPD, we restricted controls to women who reported giving at least one live birth. (*Supplementary Table S2)*

#### Exclusionary criteria for cases and controls

We excluded any case or control who met lifetime criteria for schizophrenia, schizoaffective disorder, and bipolar disorder I (including unipolar mania) *(Supplementary Table S2)*. Thus, anyone who had ICD-diagnosis of schizophrenia/psychosis, bipolar disorder, mania or reported any use of antipsychotics or lithium for psychiatric symptoms (*Supplementary Table S5*) were excluded from analyses. Application of these criteria removed 2 385 MD cases and 231 controls (*Supplementary Figure S1)*.

### Genotyping, quality control, imputation

Genotype data were available for 488 363 UKB participants, after a stringent quality control procedure and imputation using combined reference panels of Haplotype Reference Consortium (HRC) and UK10K merged with 1000 Genomes phase 3.^15^ 459 590 individuals remained after the exclusion of subjects with low-quality genotype data, without both genotype and phenotype data, consent withdrawal, and non-European ancestry. Ancestry outliers were determined based on Price et al. (2006)^20^ with a threshold of three standard deviation from the mean. (*Supplementary Figure S1)*.

### Statistical analysis

#### Genome-wide association studies (GWAS)

We generated GWAS summary statistics for MD subtypes to estimate SNP-heritability (*h*^*2*^_*SNP*_) and genetic correlations for computational efficiency. In the UKB, about 30% of the participants were found to be related to at least one other person in the cohort up to the 3^rd^ degree.^15^ Cryptic relatedness within sample could bias results in GWAS, while restricting to the unrelated individuals would cause a major loss of statistical power. We therefore performed the mixed linear model-based GWAS analysis (fastGWA) to retain related individuals in the UKB.^21^ We first constructed a sparse genetic relationship matrix (GRM) for all included individuals of European ancestries, and then conducted case-control GWAS for each subtype using fastGWA module in GCTA^21^, adjusting for sex, age, and the first 10 PCs (*Supplementary Methods S1*.*2)*.

For subtype-specific GWAS with genome-wide significant SNPs (p≤5×10^−8^), we identified independent genomic loci using SNP2GENE module in FUMA^22^ (details setting in *Supplementary Methods S1*.*2*); then compared our loci with the latest published MD GWAS results which consisted of samples from the Psychiatric Genomics Consortium (PGC), UKB, and 23andMe.^19^

#### SNP-Heritability

We estimated SNP-heritability (*h*^*2*^_*SNP*_) for each MD subtype using linkage disequilibrium score regression (LDSC).^23^ LDSC estimates *h*^*2*^_*SNP*_ by regressing GWAS summary statistics on LD scores estimated from a reference population (1000 genomes European samples). We report the *h*^*2*^_*SNP*_ estimates on the observed scale assuming 50:50 case-control ascertainment^24^ (***Figure 1a***; *Supplementary Methods S1*.*2*). For comparison, we also converted the estimates to the liability scale using two formulas: the standard conversion based on Lee et al. (2011)^25^, and the Yap et al. (2018)^26^ conversion which takes into account extreme phenotype selection (***Figure 1b***; *Supplementary Table S6, Figure S3)*.

**Figure 1.**
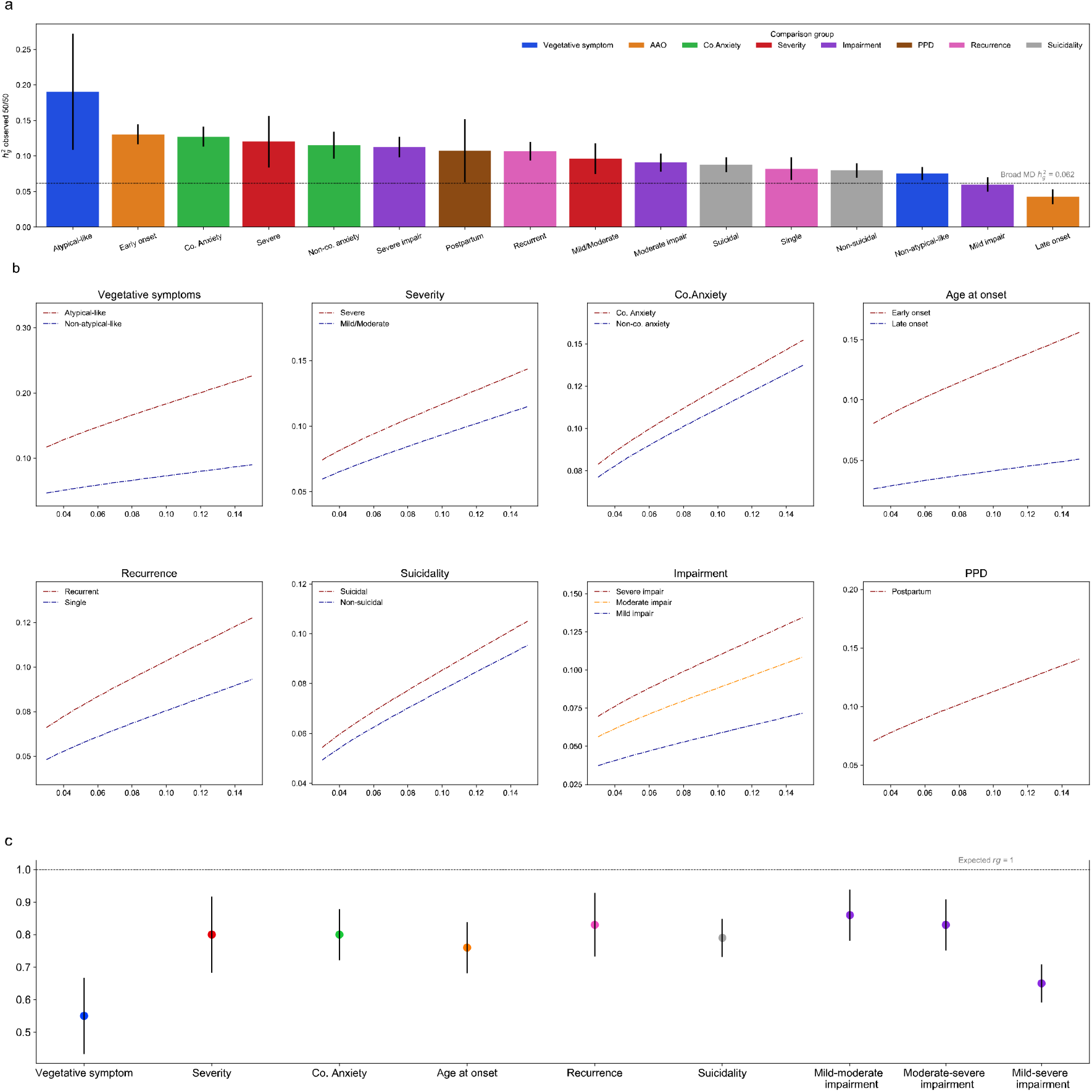
SNP-heritability and pair-wise genetic correlation for MD subtypes. **(a)** *h*^*2*^_*SNP*_ on observed scale assuming 50:50 case-control ascertainment for each MD subtype. The bars show point estimates. The error bar shows 95% CI. Same color coding is used for subtypes in the same comparison group. The horizontal line shows SNP-heritability for the broad MD phenotype (*h*^*2*^_*SNP*_=0.062). **b)** *h*^*2*^_*SNP*_ of MD subtypes on the liability-scale using Yap et al. (2018) for a range of population case prevalence. Each panel shows one comparison group. Shaded areas show 95% CI for *h*^*2*^_*SNP*_ on liability scale. Population control prevalence is fixed for each subtype as in *Supplementary Table S6*. **(c)** Pair-wise genetic correlation between subtypes within comparison groups. Error bars show 95% CI. The horizontal line shows the expected genetic correlation between subtypes under the null hypothesis (*H*_*0*_: *r*_*g*_ = 1). Result from simulations where MD cases were randomly split into two halves (with 100 replicates) showed that the expected value of *r*_*g*_ was not significantly different from the null (mean=1.04, 95% CI= 0.98-1.10). Co-anxiety: MD with comorbid anxiety; Non-co. anxiety: MD without comorbid anxiety. Colors indicate the same comparison group as in (a).

When comparing *h*^*2*^_*SNP*_ estimates within subtype comparisons, because common controls were used, we primarily considered that estimates are significantly different when non-overlapping confidence intervals are presented. We further performed statistical tests to confirm significance by splitting controls into random subsets (*Supplementary Methods S1*.*2, Table S9*). To ensure that the potential disproportionate power gain across subtype by modelling relatedness in fastGWA did not affect our *h*^*2*^_*SNP*_ comparisons, we also estimated *h*^*2*^_*SNP*_ based on unrelated samples (*Supplementary Methods S1*.*2, Figure S4*).

#### Genetic correlation

Genetic correlations (*r*_*g*_) were estimated using High-Definition Likelihood (HDL) method which yields more precise estimates of genetic correlations than LDSC *(Supplementary Methods S1*.*2)*.^27^ We estimated *r*_*g*_ within subtype comparisons using the LD reference computed from 336 000 Genomic British individuals in the UKB.^27^ To benchmark the expected *r*_*g*_ under the null hypothesis (H_0_: *r*_*g*_ = 1) in this population, we conducted a simulation analysis where MD cases were randomly split into two halves and estimated *r*_*g*_ between those two dummy-subtypes. We repeated the analysis 100 times, and calculated the mean (*Figure legend 1*).

To examine whether the subtypes differ in their genetic overlap with other psychiatric disorders and traits, we also estimated genetic correlations between these MD subtypes and 11 traits, six psychiatric disorders, neuroticism, self-reported well-being, body mass index, and two cognitive traits (***Figure 2***) and compared results within subtype comparisons. These disorders and traits were chosen given the strong evidence for their genetic correlations with MD, or in some cases, for their causal effects on MD.^13, 19^ We have used the summary statistics from the latest published GWAS for the calculations of *r*_*g*_ using HDL.^19, 28-37^

**Figure 2.**
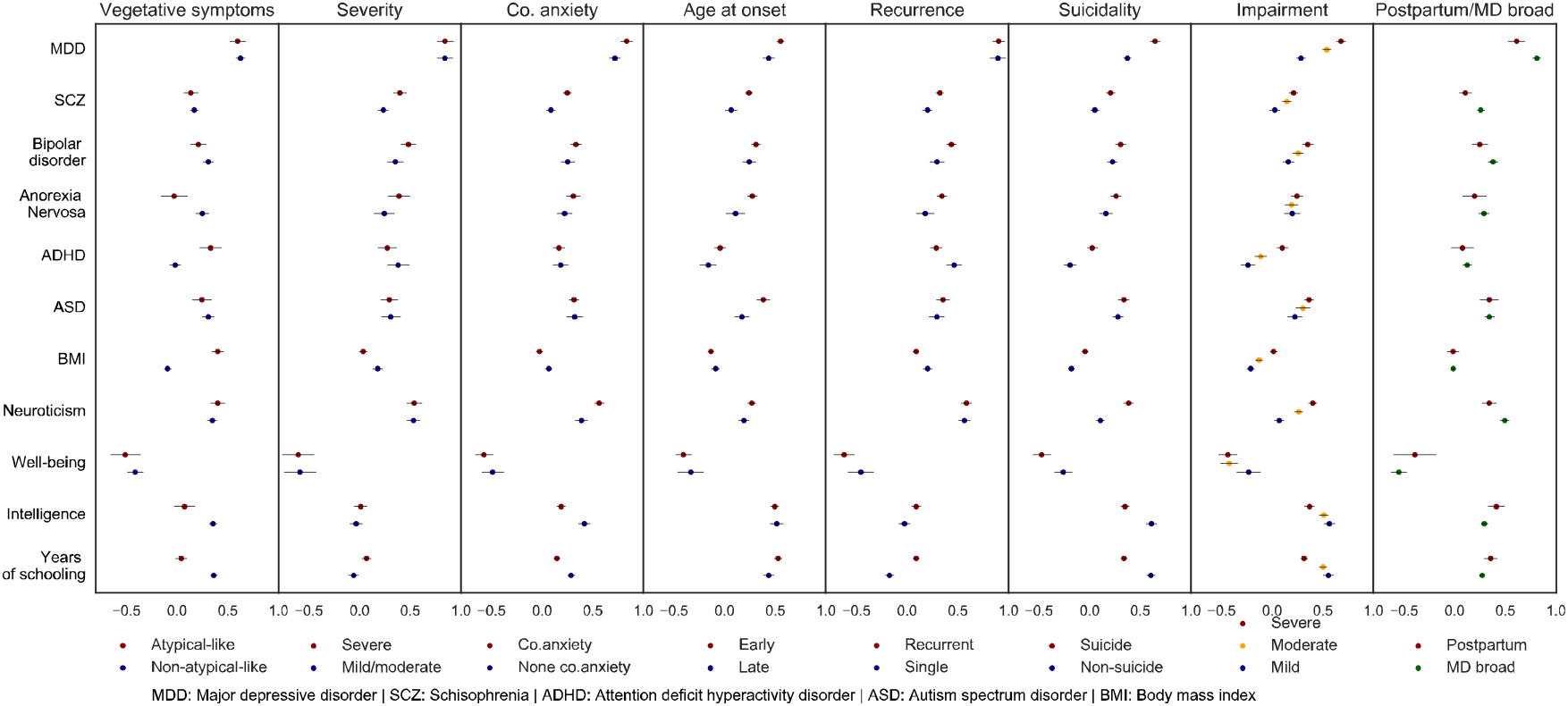
Genetic correlations (*r*_*g*_) between MD subtypes with other psychiatric disorders and related traits. Each panel shows *r*_*g*_ with other traits for each subtype comparison. Last panel shows the comparison between postpartum depression and broad MD. *r*_*g*_ with other traits for each subtype are in different colors. Error bars show 95% CI. Vertical dash lines in each panel at *r*_*g=*_0. Horizontal dash line separates psychiatric and other traits. *Co-anxiety*: MD with comorbid anxiety; *Non-co. anxiety*: MD without comorbid anxiety.

#### Sensitivity analyses

To examine whether our broad MD definition that included less strictly defined cases may bias results, we further restricted the analyses to the CIDI-based definition—previously suggested as the closest to the gold standard for diagnosing MD in the UKB^17, 38^—and performed similar analyses for all subtypes except impairment (*Supplementary Methods S1*.*2*).

## RESULTS

Of 459 590 individuals included in this study (54% females, mean age at recruitment 57 (SD 8.00)), 126 506 (27.53%) met at least one of the five definitions for MD (*i*.*e*., broad MD phenotype). After applying exclusion criteria, we retained 124 121 cases and 250 229 controls. Compared with controls, MD cases had more females (64% vs 47%), higher Townsend deprivation index which measures material deprivation with a higher score implies a greater degree of deprivation (mean −1.33, SD 3.02 vs −x1.63, SD 2.90), more lifetime smokers (57% vs 52%), but did not differ in mean BMI (mean 27.3, SD 4.6 vs 27.3, SD 5.0).

The estimates of *h*^*2*^_*SNP*_ varied across the five MD case definitions, and for the broad MD phenotype it was 6.18% (95% CI= 5.65-6.71%) on the observed scale assuming 50:50 case-control ascertainment.

### Differences in genetic components reflect subtype heterogeneity

Overall, *h*^*2*^_*SNP*_ estimates tended to be higher in MD subtypes with more severe manifestation (e.g., MD with atypical-like features, recurrent, PPD, severe impairment and severe symptoms subtypes) (***Figure 1a***). All of the subtype comparisons had higher *h*^*2*^_*SNP*_ estimates for the more severe manifestation, and three (vegetative symptoms, age at onset, and impairment) showed significant differences in *h*^*2*^_*SNP*_ estimates (***Figure 1a-b***, *Supplementary Table S9*). All examined genetic correlations within comparisons were significantly less than one and the estimates ranged between 0.55-0.86 (***Figure 1c***; *pairwise phenotypic and genetic correlation in Supplementary Figure S5*).

The *h*^*2*^_*SNP*_ estimate for MD with atypical-like features was the highest among all subtypes, and it was more than twice as high as the estimate for MD without atypical-like features with a non-overlapping 95% CI (19.04%, CI= 10.89-27.19% and 7.53%, CI=6.63-8.43%). The genetic correlation between MD subtypes with and without atypical-like features was the lowest among all comparisons (*r*_*g*_=0.55, CI=0.43-0.67) (***Figure 1c***). The two subtypes did not significantly differ in their genetic correlations with PGC MD (***Figure 2***); instead major differences were found in their correlations with anorexia nervosa and ADHD. Consistent with previous findings^14, 39, 40^, MD with atypical-like features showed a strong positive *r*_*g*_ with BMI (0.40, CI=0.34-0.46) while non-atypical-like features MD showed a small negative *r*_*g*_ instead (*r*_*g*_=-0.09, CI=-0.13 to −0.06). Furthermore, positive correlations with cognitive traits were observed in non-atypical-like features MD (*r*_*g*_=0.36 and 0.35 with educational attainment and intelligence) which were not found in MD with atypical-like features (corresponding *r*_*g*_= 0.04 and 0.07).

The MD subtype with severe symptoms had slightly higher *h*^*2*^_*SNP*_ estimate than the one with mild/moderate symptoms, although the two estimates were not significantly different. The *r*_*g*_ within comparison was significantly lower than 1 (0.80, CI=0.68-0.92). However, the two subtypes did not differ in their correlations with other traits except for a stronger *r*_*g*_ with schizophrenia found in the subtype with severe symptoms (***Figure 1-2***).

Assuming the proportions of MD cases with and without comorbid anxiety disorder at 55% and 45%, respectively^41^, the former subtype was more heritable than the latter (*h*^*2*^_*SNP*_=12.73%, CI= 11.32-14.14%, for MD with comorbid anxiety disorder, compared with 11.52%, CI=9.64-13.40%, for MD without anxiety disorder). The *r*_*g*_ within this comparison was 0.80 (CI=0.72-0.88) (***Figure 1***). Furthermore, the subtype with comorbid anxiety disorder showed higher genetic correlations with MD, schizophrenia and neuroticism, as well as lower correlations with cognitive traits, when compared with the subtype without anxiety disorder (***Figure 2***).

The *h*^*2*^_*SNP*_ of early-onset MD was three times higher than that of the late-onset subtype (13.04%, CI=11.65-14.43% compared with 4.26%, CI=3.22-5.30%). The *r*_*g*_ within comparison was 0.76 (CI=0.68-0.84). (***Figure 1***). Significant differences in their *r*_*g*_ with other traits were observed, including higher genetic correlations in early-onset MD with PGC MD, schizophrenia, anorexia nervosa, and autism spectrum disorder, than in late-onset MD (***Figure 2***).

Recurrent showed a higher *h*^*2*^_*SNP*_ estimates than single-episode MD, 10.67% (CI=9.38-11.96%) vs 8.22% (CI=6.59-9.85%). Their *r*_*g*_ was significantly lower than one (0.83, CI=0.73-0.93) *(Figure 1)*. Compared with single-episode cases, recurrent MD had stronger positive correlations with schizophrenia, bipolar disorder, anorexia nervosa, while lower genetic correlation with BMI (***Figure 2***).

The MD subtype with suicidal thoughts was slightly more heritable than the subtype without albeit the CI largely overlapped (8.79%, CI=7.75-9.83% and 7.98%, CI=6.98-8.98%). The *r*_*g*_ within this comparison was 0.79 (CI=0.73-0.85). The two subtypes in this comparison significantly differed in their genetic correlations with the majority of the other traits considered. Compared with the subtype without suicidal thoughts, the suicidal subtype showed substantially higher positive *r*_*g*_ with PGC MD, schizophrenia, neuroticism, and negative *r*_*g*_ with well-being; while its *r*_*g*_ with cognitive traits was much weaker (***Figure 1-2***).

For subtypes based on impairment, the *h*^*2*^_*SNP*_ estimates increased with the degree of impairment, roughly in a dose-response relationship, *i*.*e*., mild impairment had the lowest *h*^*2*^_*SNP*_ (6.00%, CI=4.98-7.02%), followed by moderate (9.08%, CI=7.79-10.37%) and severe impairment (11.27%, CI=9.84-12.70%). This dose-response relationship was also reflected in the pair-wise genetic correlation estimates, with the *r*_*g*_ comparing mild and severe impairment (0.65, CI=0.59-0.71) markedly lower than the other two correlations (***Figure 1***). We observed a clear trend, that is, the more severe impairment in the subtype, the stronger genetic correlation it had with other psychiatric disorders and neuroticism (positive *r*_*g*_), and with self-reported well-being (negative *r*_*g*_), while less severe impairment was more strongly associated with cognitive traits (positive *r*_*g*_) and with BMI (negative *r*_*g*_) (***Figure 2***).

The *h*^*2*^_*SNP*_ of PPD was estimated at 10.73% (CI=6.28-15.18%) which was higher compared with *h*^*2*^_*SNP*_ of broad MD phenotype. PPD showed significant positive *r*_*g*_ with other psychiatric disorders, with the strongest *r*_*g*_ observed in PGC MD as expected (0.61, CI=0.53-0.69), and with neuroticism (*r*_*g*_=0.34) and cognitive traits (*r*_*g*_=0.35 and 0.41 with educational attainment and intelligence), and a negative *r*_*g*_ with well-being (*r*_*g*_*=*-0.39) (***Figure 2***).

The broad MD definition was used above to allow sufficient statistical power in analyzing each subtype. We further assessed the impact of MD definition by performing a sensitivity analysis based on more strictly defined MD cases. The *h*^*2*^_*SNP*_ of the CIDI-based definition was 13.12%, CI=11.12-15.12% (*Supplementary Figure S2*). Restricting the analyses to the CIDI-based cases, the results were highly similar, except for the comparisons of symptom severity and recurrence, where the CIs of the *r*_*g*_ estimates now included one due to markedly reduced sample sizes in these subtypes (*Supplementary Table S8*).

### Stratified GWAS reveal novel subtype-specific loci

Over all 16 subtype-specific GWAS, we identified 47 genome-wide significant loci (45 non-overlapping) associated with nine subtypes. Less than half (22 loci) were significant in our largest GWAS of broad MD. Comparing with the latest published MD GWAS^19^, we found 14 loci that have not been reported on MD, with 3 for early-onset, 3 for recurrent, 3 for suicidal MD, 2 for non-suicidal, 1 for non-atypical-like features, 1 for moderate impairment and 1 for PPD (***Table 2***; full results on the 45 loci in *Supplementary Table S7*). The majority (64%) of these 14 loci showed no statistically significant association with the other subtype in comparison (*P*>0.05; *Supplementary Table S7*), suggesting subtype-specific effects. The chromosome 2 locus for recurrent MD, with the leading SNP rs6431690, was significant even after the stringent Bonferroni correction (*P*<3.125*10^−9^).

**Table 2.**
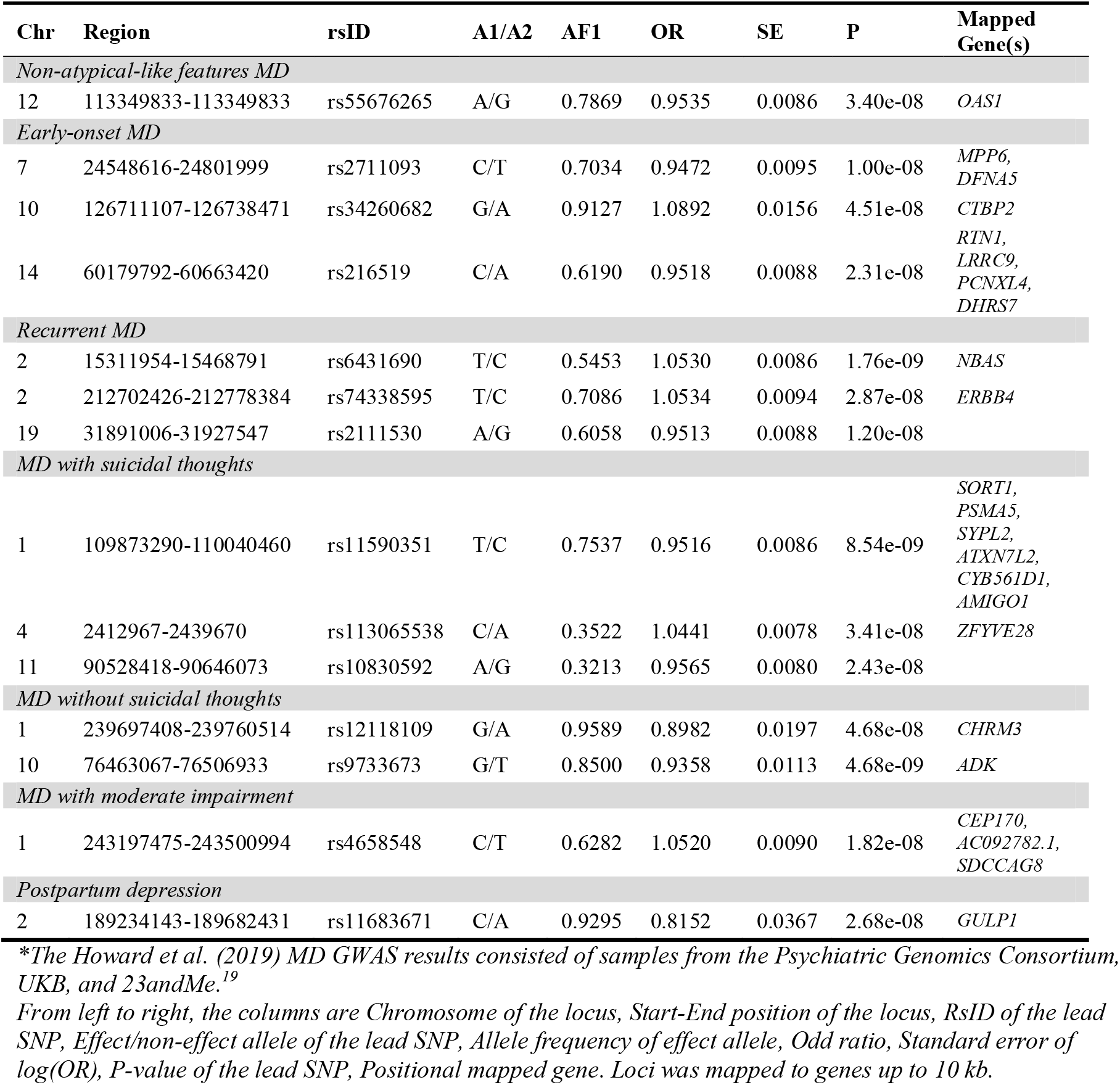
14 genome-wide significant loci from MD subtype-specific GWAS, undetected in the Howard *et al*. 2019.

## DISCUSSION

In this comprehensive report using the large-scale UKB data, we compared the genetic components of 16 MD subtypes and demonstrated that these subtypes were divergent in their *h*^*2*^_*SNP*_ and genetic correlations both within subtype comparisons and with other related disorders/traits. Our results provide convincing evidence for genetic heterogeneity within MD, as indexed by its clinical subtypes. These findings suggest that the complexity in the phenotype of MD can be effectively reduced by studying the subtypes which share partially distinct etiologies. In particular, we note the following key findings:

First, clinically-informed subtypes are, in general, genetically more homogeneous than considering all types of MD together. Accurately identifying more homogenous forms is the first step to reduce heterogeneity in MD. The majority of the subtypes showed higher estimates of *h*^*2*^_*SNP*_ compared with MD of all forms. Our results corroborated previous findings from family-based studies that early-onset, recurrent MD and PPD represent more heritable MD subtypes.^10, 12^ We further extended the list to include almost all subtypes based on our eight investigated clinical indices. Among those, MD with atypical-like features, severe episode, MD with or without comorbid anxiety disorder, and with severe impairment showed considerably higher heritability. In contrast, subtypes with lower heritability than all-form MD are those with mild impairment or with late onset.

Second, we demonstrated subtype heterogeneity in both *h*^*2*^_*SNP*_ and genetic correlations. All subtype comparisons showed non-identical genetic sharing (*i*.*e*., *r*_*g*_ between subtypes significantly differ from unity) and some had heritability differences (*i*.*e*., *h*^*2*^_*SNP*_ significantly differ between subtypes). Interestingly, the subtype comparisons on vegetative symptoms, age at onset, and impairment showed the strongest evidence for genetic heterogeneity, meaning these clinical features characterize major etiological differences within MD.

However, the observed genetic correlations across subtype comparisons were moderate to high, 0.55-0.86, revealing substantial genetic overlaps between subtypes. The level of genetic correlation can be translated into the proportion of genetic variance in one trait attributable to that of another (*r*_*g*_^2^).^17^ Thus, it would suggest about 30-70% of genetic variances are shared within subtype comparisons. In line with previous estimates of genetic correlations between male versus female MD^9^ and across MD symptoms^42^, our findings confirm that the genetic profiles of MD subtypes are only partially distinct.

The estimates of genetic correlations between subtypes need to be benchmarked against genetic correlations between different psychiatric disorders (e.g., schizophrenia and bipolar disorder, two clinically distinct psychiatric disorders, had a *r*_*g*_ of ∼0.70^31^), between different datasets but with same phenotype (e.g., mean *r*_*g*_∼0.76 across the seven cohorts at PGC MD^13^), and between different populations (e.g., *r*_*g*_∼1 between schizophrenia samples of East Asian and European ancestries^43^). Genetic correlations can be found lower than one due to differences in phenotype definitions, populations, or technical factors^44^. In this study, we minimized these potential differences by using the single large sample from UKB. We also restricted the estimation of genetic correlations to within subtype comparisons instead of pair-wise comparisons across all subtypes, to limit the impact of phenotypic differences between subtypes (*e*.*g*., we found mean *r*_*g*_ across all subtypes was indeed lower than that within comparison groups). Our genetic correlation estimates are thus reliable for quantifying overall genetic sharing between MD subtypes.

Third, the MD subtypes preserve the overall pattern of genetic sharing found between MD (of all forms) and other psychiatric disorders, but differ in the relationships with other traits. MD was shown to be positively correlated with many psychiatric disorders (e.g., *r*_*g*_∼0.3 with schizophrenia and bipolar disorders) and with BMI (*r*_*g*_=0.09), and negatively correlated with educational attainment (*r*_*g*_=-0.13).^13, 19^ A similar level of genetic correlations was found between MD subtypes and other psychiatric disorders; notably, we found stronger correlations in the MD subtypes that are more clinically challenging, especially early-onset, recurrent, suicidal, more severely impaired. Regarding their relationships with other traits, MD subtypes showed some differences compared with the broad MD phenotype. The positive correlation found between MD and BMI was only detected in MD with atypical-like features, but with a markedly higher estimate (*r*_*g*_∼0.4) than the estimate based on broad MD phenotype. This result concurred with previous findings mainly using PRS or other samples.^14, 39, 40, 45^ In contrast with the negative value found in the broad MD, we found positive correlations with educational attainment in many MD subtypes. However, this finding might be specific to the UKB cohort as previous research have shown that participation in mental health survey and other optional components is genetically correlated with higher education and intelligence.^46^

Taken together, our findings provide an improved understanding on heritable MD subtypes and overall genetic sharing between subtypes. These results have strong implications in the gene mapping strategies for MD. Current efforts predominantly aim to maximize samples size. The alternative strategy—to reduce phenotypic heterogeneity through more homogeneous phenotype—has not been fully evaluated, potentially due to theoretical and methodological challenges.^47^ This strategy relies on the premise that “clinical heterogeneity in MD emerges from an aggregation of different underlying liabilities expressed through partially distinct biological pathways”^47^ which, to the best knowledge, was not proven. Limited by a lack of large-scale dataset with deep phenotyping, prior studies were only able to focus on a few key subtypes.^5, 47^ Our comprehensive report, by contrast, convincingly demonstrated genetic heterogeneity in MD, and thus forms a strong theoretical basis for this strategy. We further illustrated the potential of such strategy by performing stratified GWAS on each subtype. This yielded the identification of 47 independent genomic loci, a third of which were undetected in the latest MD GWAS with about 5- to 10-fold more cases than in our subtype-specific analyses. These results warrant further replications in large biobanks with consistent genotyping and phenotyping. Future data collections in MD may benefit from assessing key clinical characteristics and utilizing them to reduce MD heterogeneity.

Here we used the UKB data which provide the unique opportunity to evaluate multiple subtypes with sufficient statistical power. We, however, note the following limitations in the context of interpreting the results. First, we were unable to study all MD subtypes, especially rare subtypes like psychotic, seasonal, treatment-related subtypes, as more refined clinical and treatment data would be required. We also acknowledge that the quality of phenotypic definitions varied across the subtypes studied, with those relying on self-reported and retrospective recalls of symptoms more compromised than the others. Together with the varying prevalence and underlying genetic architectures, statistical power varied across subtypes and the power gain using fastGWA may not be proportional to subtypes. “Healthy volunteer bias” was known for UKB^48^ and likely to contribute to part of our results. Finally, we used theory-driven subtyping approach in this study. New methods using data-driven approaches might hold great promises for novel subtype identification and validation.

Etiological heterogeneity hinders treatment efficacy. Our finding of ubiquitous subtype heterogeneity within MD underscores the potential of drug development and treatment optimization for patient subgroups to achieve precision psychiatry.

## Data Availability

Data used for this manuscript is not available for public access. Please contact the corresponding author for questions regarding data.
Code that was used to generate the results is available on GitHub link below.

https://github.com/Thuy-Dung-Nguyen/MD-subtypes

## URLs

Full protocol and scripts available via Github: https://github.com/Thuy-Dung-Nguyen/MD-subtypes;

UK Biobank Showcase User Guide (2017):

http://biobank.ctsu.ox.ac.uk/crystal/crystal/exinfo/ShowcaseUserGuide.pdf;

UK Biobank-Mental health web-based questionnaire (2017):

http://biobank.ctsu.ox.ac.uk/crystal/crystal/docs/mental_health_online.pdf;

GCTA-fastGWA: https://cnsgenomics.com/software/gcta/#fastGWA;

FUMA: https://fuma.ctglab.nl;

LDSC: https://github.com/bulik/ldsc;

HDL: https://github.com/zhenin/HDL;

Howard et al. 2019 MD GWAS summary results:

https://datashare.is.ed.ac.uk/handle/10283/3203.

## Acknowledgement

This research has been conducted using the UK Biobank Resource under Application Number 22224. This study was funded by the US NIMH grant (R01 MH123724) and the European Union’s Horizon 2020 research and innovation program under grant agreement No 847776. KK was supported by the University of Manitoba and US NIMH (R01 MH123724). PFS was supported by the Swedish Research Council (Vetenskapsrådet, award D0886501), the Horizon 2020 Program of the European Union (COSYN, RIA grant agreement n° 610307), and US NIMH (U01 MH109528 and R01 MH077139). YL is in part supported by a 2018 NARSAD Young Investigator Grant from the Brain & Behaviour Research Foundation and US NIMH (R01 MH123724).

The computations were enabled by resources provided by the Swedish National Infrastructure for Computing (SNIC) at UPPMAX server partially funded by the Swedish Research Council through grant agreement no. 2018-05973.

## Contributions

YL conceived the idea, designed and supervised the implementation of the study, interpreted the results, revised the manuscript. TDN conducted the analyses, interpreted the results, drafted, and revised the manuscript. AH, YX contributed to the analyses, and solving technical issues. KK, NC, RKH, CD, SH contributed to the study design, discussion on methodological issues and interpretation of the results. SH provided access to the data. PFS contributed to the discussions on methods, interpretation of the results and revising the manuscript. All authors discussed and commented on the manuscript.

