## Supplementary methods, tables and figures for "Genetic heterogeneity and subtypes of major depression"

Data supplement for

### I. SUPPLEMENTARY METHODS

#### S1.1 Materials

#### UKB cohort

All UKB participants were invited to one of the assessment centers and answered touchscreen questionnaires that contains a section about mental health, followed by an interview with a research nurse where more details about mental illness and medication use were obtained. Psychiatric disorders diagnosed by clinicians were obtained from hospital admission records. Additionally, ~158,000 participants completed a web-based mental health questionnaire (MHQ) which is based on the Composite International Diagnostic Interview (CIDI).

#### MD case definitions

The broad MD phenotype in this study is a composition of five MD definitions, including:

(1) Lifetime depression based on the CIDI Short Form, i.e., lifetime depression according to the DSM-V criteria;
(2) ICD-coded depression, i.e., any primary or secondary clinical diagnosis of major depressive disorder from linked hospital admission records;
(3) Probable MD, i.e., presence of low mood or anhedonia for ≥2 weeks and 'help-seeking';
(4) Self-reported MD, i.e., those self-reported having MD during nurse interview or mental health problems ever diagnosed by a professional as part of MHQ;
(5) MD cardinal symptoms, i.e., ever had prolonged loss of interest in normal activities; or prolonged feelings of sadness or depression.

See *Supplementary table S2* about how to derive the MD definitions from UKB data.

#### MD subtypes

See *Supplementary table S4* for methods to derive MD subtypes in UKB data.

<u>MD with atypical-like features versus non-atypical-like features.</u> Symptoms of MD were extracted from the MHQ online follow-up, and used to determine if the participant had atypical-like features versus without atypical-like features. As in a previous study[1, 2], we defined MD with atypical-like features subtype as MD cases who reported both hypersomnia (UKB field: 20534) and weight gain (UKB field: 20536), and the others were non-atypical-like features MD subtype. This definition should be considered separately from the 'atypical MD', which requires the endorsement of full DSM criteria including mood reactivity and other atypical symptoms.

<u>Severe versus mild/moderate MD</u>. The subtypes on symptom severity compared the cases with mild/moderate versus severe symptoms. Mild/moderate MD included those with probable recurrent MD (moderate) defined by Smith et al.[3] (UKB field: 20126), and/or ICD-diagnoses of mild (F320, F330) or moderate depression episode (F321, F331) (UKB field: 41202, 41204). Severe MD included probable recurrent MD (severe) defined by Smith et al.[3] (UKB field: 20126); and/or ICD-diagnoses of severe MD (F322, F323, F332, F333) (UKB field: 41202, 41204).

<u>MD with or without comorbid anxiety disorder.</u> The MD subtypes with or without comorbid anxiety disorder were defined using both self-reported and ICD-coded diagnoses. Participants of the MHQ reported mental health problems diagnosed by a professional (UKB 20544). Among the MD cases, we defined the subtype with comorbid anxiety disorder as those who also reported social anxiety/phobia, panic attacks, and anxiety, nerves/generalized anxiety disorder in the MHQ, and/or those with ICD diagnoses (UKB field: 41202, 41204) of anxiety disorder (F40, F41). For comparison, MD without comorbid anxiety disorder included MD cases with neither self-reported nor ICD-codes anxiety disorder.

Early versus late onset MD. Among >89,000 participants who answered the MHQ online follow-up, those who had MD cardinal symptoms reported the age at which they first experienced a ≥2-week episode (UKB field: 20433). Similar to previous studies[4, 5], we categorized the continuous variable of age at onset into octiles, then grouped the first 3 octiles into early-onset subtype (age onset ≤30) and the last 3 octiles into late-onset subtype (age onset ≥44).

Recurrent versus single-episode MD. Recurrent MD subtype was defined as those with probable recurrent MD defined by Smith et al. (2013)[3] (UKB field: 20126), and/or with ≥2 episodes of at least two weeks of cardinal symptoms (UKB field: 20442)[6], and/or ICD-diagnosis of recurrent MD (F33). Single-episode subtype included probable MD cases with one episode of feeling depressed (UKB field: 20126), and/or those self-reported a single episode (UKB field: 20442)[6], and/or ICD-diagnosis of non-recurrent MD (F32).

MD with or without suicidality. The subtype MD with suicidal thoughts included those who had reported thoughts of death during worst depression (UKB field: 20437); and/or those with recent thoughts of suicide or self-harm (UKB field: 20513). Non-suicidal subtype included MD cases who did not report suicidal thoughts.

MD with mild, moderate, severe impairment. Individuals who reported cardinal symptoms of MD were asked about the impact of MD on normal roles, including study/employment, childcare and housework, leisure pursuits, during the worst period of depression (UKB field: 20440). We categorized ‘not at all/a little impact’ as mild impairment, ‘somewhat’ as moderate impairment and ‘a lot’ as severe impairment.

Postpartum depression (PPD) cases were identified from three sources: women who reported post-natal depression during the nurse interview at the baseline recruitment (UKB field: 20002, code 1531); cases with MD cardinal symptoms related to childbirth (UKB field: 20445); and women with ICD diagnosis of mental and behavioral disorders associated with the puerperium (F53 in UKB fields: 41202, 41204).

#### S1.2 Statistical analysis

#### Genome-wide association studies (GWAS)

For GWAS, we constructed a genetic relationship matrix (GRM) for all UKB individuals of European ancestries based on 575,805 LD-pruned HapMap3 variants (LD-pruning parameters in PLINK 1.9: window size = 1Mb, step size = 100, $r^2$ = 0.9). From a full GRM, we derived a sparse GRM by setting all elements <0.05 to 0 (--grm-sparse flag in GCTA). We then conducted case-control GWAS for each subtype using fastGWA[2], adjusting for sex, age, and the first 10 PCs. Summary statistics were available for ~9.7 million common variants in each GWAS after filtering out those with low imputation quality (INFO<0.6). Compared to other methods, fastGWA has efficiency as the advantage. Since we have a large sample size and 30% of the sample is related, using fastGWA is more computationally efficient and help maximizing power by retaining related individuals. Simulation showed that results obtained from fastGWA was nearly identical to that from other methods like REML algorithm implemented in GCTA22 or test statistic computed using the GRAMMAR-GAMMA approximation.[2]

For annotation in FUMA[7], we applied the following parameters for lead SNPs: maximum P-value of lead SNPs < 5e-8, r2 threshold to define independent significant SNPs >=0.6, 2nd r2 threshold to define lead SNPs >=0.1, maximum distance between LD blocks to merge into a locus <250kb, with 1000G Phase3 European ancestry as reference panel. The mapped genes are based on positional and expression quantitative trait loci (eQTL).

**SNP-Heritability**

We used linkage disequilibrium score regression (LDSC)[8] to estimate SNP-heritability ($h^2_{SNP}$) on observed scale based on 50:50 case-control ascertainment[9] for each MD subtype. LDSC estimates SNP-heritability by regressing GWAS summary statistics on LD scores estimated from a reference population. We used LD scores from the European samples in the 1000 genomes reference panel. For each subtype, we calculated N effective using the formula N_effective =4*p*(1-p)*N, with sample case proportion p and total sample size N.

For comparison, we estimated heritability on observed scale (not specifying N_case/N_control in LDSC software), then converted into liability scale using two methods. First, we converted $h^2_{SNP}$ from the observed to the liability scale based on the standard conversion from Lee et al (2011)[10] (Equation 23), which takes into account sample prevalence and population prevalence. Second, we corrected for oversampling and extreme phenotype selection using sample prevalence, population case and control prevalence based on Yap et al. (2018) - formula 1- *Supplementary method*[11]. The main *Figure 1b* presents $h^2_{SNP}$ on liability scale with a range of population case prevalence for each subtype based on Yap et al. (2018). Results from the 3 methods are presented in *Supplementary table S6* and *figure S3*.

We confirmed significant differences in $h^2_{SNP}$ estimates within subtype comparisons by splitting controls into 2 or 3 random subsets. Each control subset was combined with subtype cases for GWAS and estimating $h^2_{SNP}$ (*Supplementary table S9*).

To ensure that the potential disproportionate power gain across subtype by modelling relatedness in fastGWA did not affect our heritability comparisons, we also estimated $h^2_{SNP}$ based on unrelated samples. After removing all genetic-inferred related individuals[12], we conducted GWAS using logistic regression in Plink, then estimated $h^2_{SNP}$ using LDSC. (*Supplementary Figure S4*).

**Genetic correlation**

Genetic correlations ($r_g$) were estimated using a recent extension of LDSC, High-Definition Likelihood (HDL) method.[13] Similar principle as in estimating heritability, the genetic correlation between two traits is estimated by contrasting the covariance matrix of z-scores 1 (trait 1)-z-scores 2 (trait 2) and the LD scores matrix.[14] HDL exploits all elements of the z-scores covariance matrix instead of just the diagonal elements as in LDSC, and thus yields more precise estimates of genetic correlations compared to LDSC.[13] We estimated genetic correlations within subtype comparisons, using the LD reference computed from 336 000 Genomic British individuals in the UKB.

For the primary analysis, we estimated genetic correlation for each pair of subgroups within a comparison group. GWAS summary statistics data were processed into the right format with z-scores by HDL.data.wrangling.R function in HDL program (https://github.com/zhenin/HDL/wiki/Format-of-summary-statistics). LD reference panel used is the eigenvalues and eigenvectors of LD matrices for the European-ancestry population. The matrices were computed from 335,265 Genomic British individuals in UK Biobank (https://github.com/zhenin/HDL/wiki/Reference-panels). Details about the HDL program could be found at https://github.com/zhenin/HDL/wiki.

**Sensitivity analyses**

It has been argued that results relying on minimal phenotyping (*i.e.*, the definitions of MD are not strictly based on structured clinical interviews, but instead rely on a few self-reported answers) may bias views of genetic architecture of MD.[6] To examine whether our broad MD definition may bias the results, we further restricted the analyses to the CIDI-based definition—previously suggested as the closest to the gold standard for diagnosing MD in the UKB[6, 15]—and estimated $h^2_{SNP}$ and genetic correlations for all subtypes except impairment because moderate to severe impairment is part of the CIDI-based MD criteria.

Cases of subtypes were defined using the same criteria as primary analyses, then we only included people who also met criteria for CIDI-based MD *(Supplementary table S4).* Control group was the same as in the primary GWAS analyses. We estimated $h^2_{\text{SNP}}$ on the observed scale based on 50:50 case-control ascertainment[9], and on liability scale using conversion in Yap et al. (2018)[11] *(Supplementary table S6).* Estimates of $h^2_{SNP}$ and $r_g$ are presented in *Supplementary table S8*.

In the primary analyses, we considered that subtype heritability within the same comparison group are significantly different when non-overlapping confidence intervals are presented, we performed statistical tests to confirm significance for the three subtype categories with non-overlapped CI from the original results (age at onset, suicidality, and impairment). We split the control group into 2 random subsets, each subtype in the same category consisted of one control subset. We conducted GWAS using fastGWA and estimated heritability with LDSC. Results are presented in *Supplementary table S9*.

### II. SUPPLEMENTARY TABLES

#### Table S1 Summary of current literature on MD subtype heterogeneity

| Subtype category | Clinical features[†] | Comparisons[†] | Heritability | | Genetic overlap | | Subtype specific GWAS |
|---|---|---|---|---|---|---|---|
| | | | $h^2$ | $h^2_{SNP}$ | $r_g$ within comparison | Overlap w other traits (PRS/$r_g$) | |
| Symptom-based | **Vegetative symptom** | **Atypical**<br>**Typical** | | Null[††16-18]<br>Null[††16-18] | 0.82 (0.33-1.31)[18]<br>0.54 (0.27-0.81)[16] | PRS of metabolic traits more associated with atypical MD[16, 17]<br>$r_g$ atypical w. BMI= 0.53 (95% CI= 0.23-0.82)[18]<br>$r_g$ typical w. BMI= -0.28 (95% CI=-0.55;-0.01)[18] | |
| | **Symptom severity** | **Severe**<br>**Mild/ moderate** | | | | | |
| | **Comorbid anxiety disorder** | **With anxiety**<br>**Without anxiety** | | | | | |
| Time of onset/course | **Age at onset** | **Early**<br>**Late** | 0.21(0.12-0.28)[19]<br>0.23(0.13-0.32)[19] | | 0.85 (0.66-0.98)[19] | Stronger PRS association between early-onset MDD with BIP/SCZ[4] | 1 locus[4] |
| | **Recurrence** | **Single**<br>**Recurrent** | 0.28(0.14-0.41)[19]<br>0.41(0.2-0.6)[19] | | 0.86 (0.68-0.97)[19] | | 2 loci in Han Chinese[20] |
| Treatment-related & other outcomes | **Suicidality** | **w suicidal thoughts**<br>**w/o suicidal thoughts** | | | | | |
| | **Impairment** | **Mild**<br>**Moderate**<br>**Severe** | | | | | |
| | **PPD** | **PPD** | 0.44(0.35-0.52)[21] | | | | |
| Etiologically-based | Environment | With trauma/ adversity<br>Without trauma/ adversity | | Inconsistent[†††22, 23]<br>Inconsistent [†††22, 23] | 0.77 (0.48-1.05)[22]<br>0.64 (0.19-1.09)[23] | Stronger association between MD w trauma and body composition, socioeconomic[22] | Not found[23]<br>3 loci in Han Chinese[23] |
| Gender-based | Sex | Female<br>Male | 0.44(0.25-0.61)[19]<br>0.35(0.08-0.63)[19] | 0.22(0.06)[24]<br>0.18(0.06)[24] | 0.89 (0.87-0.91)[25] | | Not found[24]<br>1 locus[24] |

*MD: major depression. PPD: postpartum depression. BIP: Bipolar disorder. SCZ: Schizophrenia.* $h^2$: heritability. $h^2_{SNP}$: SNP-heritability. $r_g$: Genetic correlation. PRS: Polygenic Risk Score

[†] The subtypes examined in this study were highlighted in bold. Subtypes with or without trauma were not studied here because similar analyses using the UK biobank were available; and sex-specific subtypes were not studied because it has been heavily studied and evidence were convincing.

†† Compared with MD with typical symptoms, $h^2_g$ estimates for MD with atypical symptoms was not significantly different in Milaneschi et al. (2016)[17], 0.43 (95% CI=0.04-0.82) vs 0.38 (95% CI=0.05-0.71); in Milaneschi at al. (2017)[18], 0.11 (95% CI=0.05-0.17) vs 0.11 (95% CI=0.07-0.15); and in Badini et al. (2020)[16], 0.14 (95% CI=0.03-0.15) vs 0.12 (95% CI=0.08–0.16).

††† Compared with MD without trauma, $h^2_g$ estimate was higher for MD with trauma in Coleman et al. (2020)[22], 0.24 (95% CI=0.18-0.31) vs 0.12 (95% CI=0.07-0.16), but lower in Peterson et al. (2018)[23], 0.34 (95% CI=0.03-0.65) vs 0.38 (95% CI=0.29-0.47).

### Table S2 Method to derive of MD cases, control and exclusion criteria in UKB

| Measure | Description | Criteria | Derive method |
|---|---|---|---|
| CIDI-SF based MD | Lifetime or current MDD according to DSM-V criteria | **Life-time CIDI MD**<br>5/8 criteria A1-A9 (lack A5) with at least one criteria A1 or A2 plus criteria BCDE as below:<br>A1 (UKB 20446)<br>A2 (UKB 20441)<br>A3 (UKB 20536)<br>A4 (UKB 20532)<br>A6 (UKB 20449)<br>A7 (UKB 20450)<br>A8 (UKB 20435)<br>A9 (UKB 20437)<br>B - Impact on normal roles during worst period of depression (UKB 20440)<br>C - Not substance abuse (UKB 41202 + 41204)<br>D- Not SCZ or psychotic (UKB 41202 + 41204<br>E - Not Maniac (UKB 41202 + 41204)<br>**Current CIDI MD**<br>- 5/9 criteria with at least one criteria A1 or A2 (each satisfied A criteria scores 1)<br>A1 (UKB 20510) = 4<br>A2 (UKB 20514) = 4<br>A3 (UKB 20511) = 4<br>A4 (UKB 20517) = 4<br>A5 (UKB 20518) = 4<br>A6 (UKB 20519) = 4<br>A7 (UKB 20507) = 4<br>A8 (UKB 20508) = 4<br>A9 (UKB 20513) = 4<br>B - Plus Impact on normal roles during worst period of depression | **Life-time CIDI MD**<br>UKB20446 = 1 or UKB20441 = 1<br>and<br>sum UKB(20446, 20441, 20536, 20532, 20449, 20450, 20435, 20437) >=5<br>and<br>UKB20440 = 3<br>and<br>UKB41202 != (F10-F19, F20-F29, F30)<br>and<br>UKB41204 != (F10-F19, F20-F29, F30)<br>**Current CIDI MD**<br>UKB20510 = 1 or UKB20514 = 1<br>and<br>sum UKB(20510, 20514, 20511, 20517, 20518, 20519, 20507, 20508, 20513) >=5<br>and<br>UKB20440 = 3<br>and<br>UKB41202 != (F10-F19, F20-F29, F30)<br>and<br>UKB41204 != (F10-F19, F20-F29, F30)<br>**CIDI-based MD**<br>Life-time CIDI MD = 1<br>or Current CIDI MDD = 1 |

| | | | |
|---|---|---|---|
| | | (UKB 20440)<br>C - Plus Not substance abuse (UKB 41202 + 41204)<br>D - Plus Not SCZ or psychotic (UKB 41202 + 41204)<br>E - Plus Not Maniac (UKB 41202 + 41204) | |
| ICD-coded MD | Clinician diagnosis of MDD | Any primary or secondary diagnosis of a depressive mood disorder from linked hospital admission records (UKB 41202 + 41204) | UKB41202 = F32/F33/F34/F38/F39<br>or<br>UKB41204 = F32/F33/F34/F38/F39 |
| Probable MD | | Depressed/down for a whole week (UKB: 4598);<br>And<br>At least 2 weeks duration (UKB: 4609);<br>And<br>Ever seen a GP or psychiatrist for nerves, anxiety or depression" (UKB 2090 + 2010)<br>Or<br>Ever anhedonia for a whole week (UKB 4631);<br>plus<br>At least 2 weeks duration (UKB 5375);<br>And<br>Ever seen a GP or psychiatrist for nerves, anxiety, or depression (UKB 2090 + 2010) | UKB20126 = 3/4/5 |
| Self-reported MD | Self-reported ever had MDD diagnosis or non-cancer diseases including MD | Self-reported depression (UKB 20002)<br>or<br>Self-reported Mental health problems ever diagnosed by a professional (UKB 20544) | UKB20002 = 1286<br>or<br>UKB20544 = 11 |
| MD cardinal symptoms | Ever had indication of depression | Ever had prolonged loss of interest in normal activities (UKB 20441); or prolonged feelings of sadness or depression (UKB 20446) | UKB20441 = 1<br>or<br>UKB20446 = 1 |

| | | | |
|---|---|---|---|
| MD broad phenotype | The MD broad definition to define cases in the study | Meet at least 1 measure 1-5 | CIDI-SF based MD = 1<br>or<br>ICD-coded MD = 1<br>or<br>Probable MD = 1<br>or<br>Self-reported MD = 1<br>or<br>MD cardinal symptoms = 1 |
| Help-seeking MD | Self-reported help-seeking behavior for mental health difficulties | Ever seen a general practitioner (GP) for nerves, anxiety, tension or depression (UKB 2090)<br>or<br>Ever seen a psychiatrist for nerves, anxiety, tension or depression (UKB 2010) | UKB2090 = 1<br>or<br>UKB2100 = 1 |
| Antidepressant use | Self-reported use of antidepressants as regular medication and health supplements | Reported use of any antidepressant (UKB 20003) | UKB20003 = any code in the list of antidepressants |
| Control MD | Controls of MD population | Not meeting any of 7 MD definitions 1-7 as above | CIDI-SF based MD != 1<br>and<br>ICD-coded MD != 1<br>and<br>Probable MD != 1<br>and<br>Self-reported MD != 1<br>and<br>MD cardinal symptoms != 1<br>and<br>Help-seeking MD !=1<br>and<br>Antidepressant use != 1 |
| Control postpartum MD | Controls of PPD population | Women (UKB 31)<br>and<br>Have given >= 1 live birth (UKB 2734) | Control MD = 1<br>and<br>UKB31 = 0 |

| | | | |
|---|---|---|---|
| | | and<br>Not meeting any of 7 MD definitions 1-7 as above | and<br>UKB2734 > 0 |
| Control comorbid anxiety MD | Control for cases in the comorbid anxiety subtype | Not meeting any of 7 MD definitions 1-7 as above<br>and<br>Not having anxiety disorder F40/F41 (UKB 41202, 41204) | Control MD = 1<br>and<br>UKB41202 != (F40, F41)<br>and<br>UKB41204 != (F40, F41) |
| Exclusion criteria | Criteria to excluded cases and controls | Do not have diagnosis of SCZ/psychosis (F20-F29), BP (F31) and Manias (F30) (UKB 41202 + 41204)<br>and<br>Do not used antipsychotics or lithium (UKB 20003) | UKB41202 != (F10-F19, F20-F29, F30)<br>and<br>UKB41204 != (F10-F19, F20-F29, F30)<br>and<br>UKB20003 != any code in the list of antipsychotics and lithium |

**Note: != means NOT EQUAL; coding 0 = No; 1 = Yes for binary variables; otherwise, check https://biobank.ctsu.ox.ac.uk/crystal/index.cgi*

**Table S3 MD definitions and derived subtypes**

| Source | MD definitions | N (%*) | Derived subtype |
|---|---|---|---|
| MHQ online questionnaire | CIDI-SF based MD | 17,121 (3.76%) | - Recurrence |
| Hospital admission records | ICD-coded MD | 12,135 (2.66%) | - Severity<br>- Comorbid anxiety<br>- Postpartum |
| Touchscreen questionnaire (mental health) | Probable MD | 28,628 (6.28%) | - Recurrence<br>- Severity |
| Nurse interview | Self-reported MD | 50,857 (11.15%) | - Comorbid anxiety<br>- Postpartum |
| MHQ online questionnaire | MD cardinal symptoms | 83,774 (18.37%) | - Age at onset<br>- Suicidal thought<br>- Atypical-like features<br>- Impairment |

*CIDI-SF = Composite International Diagnostic Interview Short Form | MHQ = Mental health questionnaire*
**Proportion of total 459,590 included sample*

#### Table S4 Method to derive MD subtypes in UKB data

| Subtypes | Description | Subtype within MD broad phenotype[a] | Subtype within CIDI-SF based MD[b] |
|---|---|---|---|
| **Vegetative symptoms** | | | |
| Atypical-like features | MD with atypical features Hypersomnia (UKB20534) and weight gain (UKB20536) | MD broad phenotype = 1<br>and<br>(UKB20534 = 1 and UKB20536 = 1) | CIDI-SF-based MD = 1<br>and<br>(UKB20534 = 1 and UKB20536 = 1) |
| Non-atypical-like features | MD without atypical features. i.e., did not have hypersomnia and weight gain at the same time | MD broad phenotype = 1<br>and<br>(UKB20534 = 0 and/or UKB20536 = 0) | CIDI-SF-based MD = 1<br>and<br>(UKB20534 = 0 and/or UKB20536 = 0) |
| **Severity** | | | |
| Severe | Severe episode MD ascertained by self-report and ICD diagnostic codes (UKB20126, UKB41202, UKB41204) | MD broad phenotype = 1<br>and<br>(UKB20126 = 3 or UKB41202 = F322/F323/ F332/F333 or UKB41204 = F322/F323/ F332/F333) | CIDI-SF-based MD = 1<br>and<br>(UKB20126 = 3 or UKB41202 = F322/F323/ F332/F333 or UKB41204 = F322/F323/ F332/F333) |
| Mild/moderate | Mild and moderate MD ascertained by self-report and ICD diagnostic codes (UKB20126, UKB41202, UKB41204) | MD broad phenotype = 1<br>and<br>(UKB20126 = 4 or UKB41202 = F320/F321/ F330/F331 or UKB41204 = F320/F321/F330/F331)<br>and<br>not included in severe subtype | CIDI-SF-based MD = 1<br>and<br>(UKB20126 = 4 or UKB41202 = F320/F321/ F330/F331 or UKB41204 = F320/F321/F330/F331)<br>and<br>not included in severe CIDI-SF -based subtype |
| **Comorbid anxiety** | | | |
| Comorbid anxiety | MD WITH comorbid anxiety, panic attacks, social anxiety/phobia (UKB 20544, UKB41202, UKB41204) | MD broad phenotype = 1<br>and<br>(UKB20544 = 1/6/15 or UKB41202 = F40, F41 or UKB41204 = F40, F41) | CIDI-SF-based MD = 1<br>and<br>(UKB20544 = 1/6/15 or UKB41202 = F40, F41 or UKB41204 = F40, F41) |
| Non-comorbid anxiety | MD WITHOUT comorbid anxiety, panic attacks, social anxiety/phobia (UKB 20544, UKB41202, UKB41204) | MD broad phenotype = 1<br>and<br>(UKB20544 != 1/6/15 and UKB41202 != F40, F41 and UKB41204 != F40, F41) | CIDI-SF-based MDD = 1<br>and<br>(UKB20544 != 1/6/15 and UKB41202 != F40, F41 and UKB41204 != F40, F41) |
| **Age at onset** | | | |

| | | | |
|---|---|---|---|
| Early onset | Age at first episode of depression ≤ 30 (UKB20433) | MD broad phenotype = 1<br>and<br>UKB20433 = [2,30] | CIDI-SF-based MD = 1<br>and<br>UKB20433 = [2,30] |
| Late onset | Age at first episode of depression ≥ 44 (UKB20433) | MD broad phenotype = 1<br>and<br>UKB20433 = (43,78] | CIDI-SF-based MD = 1<br>and<br>UKB20433 = (43,78] |
| **Recurrence** | | | |
| Recurrent | MD with recurrent episodes (UKB20126, UKB41202, UKB41204) | MD broad phenotype = 1<br>and<br>(UKB20126 = 3/4 or (CIDI-SF-based MDD = 1 and UKB20442 ≥ 2) or UKB41202 = F33 or UKB41204 = F33) | CIDI-SF-based MD = 1<br>and<br>(UKB20126 = 3/4 or (CIDI-SF-based MDD = 1 and UKB20442 ≥ 2) or UKB41202 = F33 or UKB41204 = F33) |
| Single | MD with single episode (UKB20126, UKB41202, UKB41204) | MD broad phenotype = 1<br>and<br>(UKB20126 = 5 or (CIDI-SF-based MDD = 1 and UKB20442 < 2) or UKB41202 = F32 or UKB41204 = F32)<br>and<br>Not in recurrent subtype | CIDI-SF-based MD = 1<br>and<br>(UKB20126 = 5 or (CIDI-SF-based MDD = 1 and UKB20442 < 2) or UKB41202 = F32 or UKB41204 = F32)<br>and<br>Not in CIDI-SF-based recurrent MD subtype |
| **Suicidality** | | | |
| Yes | Having suicidal thought during the worst period of or recent thoughts of suicide or self-harm more than half of days or nearly every day (UKB20437, UKB20513) | MD broad phenotype = 1<br>and<br>(UKB20437 = 1 or 20513 = 3/4) | CIDI-SF-based MD = 1<br>and<br>(UKB20437 = 1 or 20513 = 3/4) |
| No | Not having suicidal thought or recent thoughts of suicide or self-harm (UKB20437, UKB20513) | MD broad phenotype = 1<br>and<br>(UKB20437 = 0 and UKB20513 = 1) | CIDI-SF-based MD = 1<br>and<br>(UKB20437 = 0 and UKB20513 = 1) |
| **Impairment** | | | |
| Mild | Impact on normal roles during worst period of depression is not at all or a little (UKB20440) | MD broad phenotype = 1<br>and<br>UKB20440 = 0/1 | N/A |
| Moderate | Impact on normal roles during worst period of depression somewhat (UKB20440) | MD broad phenotype = 1<br>and<br>UKB20440 = 2 | N/A |

| | | | |
|---|---|---|---|
| Severe | Impact on normal roles during worst period of depression a lot (UKB20440) | MD broad phenotype = 1<br>and<br>UKB20440 = 3 | N/A |
| **Postpartum MD** | | | |
| MD relating to child birth | MD relating to child birth ascertained by self-report (UKB 20002, UKB 20445) and ICD code (UKB: 41202 and 41204) | MD broad phenotype = 1<br>and<br>(UKB20002 = 1531 or UKB20445 = 1 or UKB41202 = F53 or UKB41204 = F53) | CIDI-SF-based MD = 1<br>and<br>(UKB20002 = 1531 or UKB20445 = 1 or UKB41202 = F53 or UKB41204 = F53) |

*[a]Subtypes in primary analyse | [b]Subtypes in sensitivity analyses*
*!= means NOT EQUAL | coding 0 = No; 1 = Yes for binary variables; otherwise, check https://biobank.ctsu.ox.ac.uk/crystal/index.cgi*

#### Table S5 Medication list

| ID† | Medication name | ID† | Medication name |
|---|---|---|---|
| **Antidepressant** | | | |
| 1140879616 | amitriptyline | 1141151978 | reboxetine |
| 1140921600 | citalopram | 1141152736 | zispin |
| 1140879540 | fluoxetine | 1140867640 | doxepin |
| 1140867878 | sertraline | 1140867920 | moclobemide |
| 1140916282 | venlafaxine | 1140867850 | phenelzine |
| 1140909806 | dosulepin | 1140879544 | fluvoxamine |
| 1140867888 | paroxetine | 1141200570 | yentreve |
| 1141152732 | mirtazapine | 1140867934 | triptafen |
| 1141180212 | escitalopram | 1140867758 | surmontil |
| 1140879634 | trazodone | 1140867914 | tranylcypromine |
| 1140867876 | prozac | 1140867820 | allegron |
| 1140882236 | seroxat | 1141151982 | edronax |
| 1141190158 | cipralex | 1140882244 | molipaxin |
| 1141200564 | duloxetine | 1140879556 | mianserin |
| 1140867726 | lofepramine | 1140867852 | nardil |
| 1140879620 | clomipramine | 1140867860 | faverin |
| 1140867818 | nortriptyline | 1140917460 | nefazodone |
| 1140879630 | imipramine | 1140867938 | Amitriptyline + chlordiazepoxide |
| 1140879628 | dothiepin | 1140867856 | isocarboxazid |
| 1141151946 | cipramil | 1140867922 | manerix |
| 1140867948 | amitriptyline | 1140910820 | maoi |
| 1140867624 | prothiaden | 1140882312 | sinequan |
| 1140867756 | trimipramine | 1140867944 | Tranylcypromine + trifluoperazine |
| 1140867884 | lustral | 1140867784 | ludiomil |
| 1141201834 | cymbalta | 1140867812 | norval |
| 1140867690 | anafranil | 1140867668 | tryptizol |
| **Antipsychotic** | | | |
| 1140928916 | olanzapine | 1141202024 | abilify |
| 1141152848 | quetiapine | 1140882098 | fluphenazine |
| 1140867444 | risperidone | 1140867184 | haldol |
| 1140879658 | chlorpromazine | 1140867092 | serenace |
| 1140868120 | trifluoperazine | 1140882320 | clozaril |
| 1141153490 | amisulpride | 1140910358 | cpz |
| 1140867304 | sulpiride | 1140867208 | perphenazine |
| 1141152860 | seroquel | 1140909802 | levomepromazine |
| 1140867168 | haloperidol | 1140867134 | pericyazine |
| 1141195974 | aripiprazole | 1140867306 | dolmatil |
| 1140867244 | stelazine | 1140867210 | fentazin |

| | | | |
|---|---|---|---|
| 1140867152 | depixol | 1140867398 | fluphenazine |
| 1140909800 | flupentixol | 1140867078 | benperidol |
| 1140867420 | clozapine | 1140867218 | pimozide |
| 1140879746 | promazine | 1141201792 | zaponex |
| 1141177762 | risperdal | 1141200458 | denzapine |
| 1140867456 | modecate | 1140867136 | neulactil |
| 1140867952 | fluanxol | 1140879750 | thioridazine |
| 1140867150 | flupenthixol | 1140867180 | dozic |
| 1141167976 | zyprexa | 1140867546 | fluspirilene |
| 1140882100 | zuclopenthixol | 1140928260 | panadeine |
| 1140867342 | clopixol | 1140927956 | sertindole |
| 1140863416 | largactil | | |
| **Lithium** | | | |
| 1140867490 | lithium product | 1140867504 | priadel 200mg m/r tablet |
| 1140867494 | camcolit 250 tablet | 1140867518 | litarex 564mg m/r tablet |
| 1140867498 | liskonum 450mg m/r tablet | 1140867520 | li-liquid 5.4mmol/5ml oral solution |
| 1140867500 | phasal 300mg m/r tablet | | |

*This list is from Davis et al., 2019 (PMC6877131)[26] | †ID in UKB data (field 20003)*

### Table S6 Prevalence of subtypes to convert $h^2_g$ from observed to liability scale

| Phenotype | $h^2_{SNP}$ observed scale | SE observed scale | N case | N control | N effective[a] | Sample case prevalence[b] | Population case prevalence[c] | Population control prevalence[d] | Subtype proportion in literature | Source for subtype proportion | $h^2_{SNP}$ liability Yap[e] | SE liability Yap[e] | $h^2_{SNP}$ liability Lee[f] | SE liability Lee[f] | $h^2_{SNP}$ observed 50/50[g] | SE observed 50/50[g] |
|---|---|---|---|---|---|---|---|---|---|---|---|---|---|---|---|---|
| Atypical-like features | 0.0086 | 0.0019 | 2904 | 250229 | 11483 | 0.0115 | 0.0450 | 0.85 | 0.30 | PMID: 29033570 | 0.1335 | 0.0295 | 0.1555 | 0.0344 | 0.1904 | 0.0416 |
| Non-atypical-like features | 0.0400 | 0.0024 | 46900 | 250229 | 157988 | 0.1578 | 0.1050 | 0.85 | 0.70 | | 0.0748 | 0.0045 | 0.0804 | 0.0048 | 0.0753 | 0.0046 |
| Severe | 0.0143 | 0.0022 | 7923 | 250229 | 30719 | 0.0307 | 0.0750 | 0.85 | 0.50 | PMID: 29450462 | 0.1028 | 0.0158 | 0.1154 | 0.0178 | 0.1202 | 0.0184 |
| Mild/ Moderate | 0.0159 | 0.0018 | 11300 | 250229 | 43247 | 0.0432 | 0.0750 | 0.85 | 0.50 | | 0.0823 | 0.0093 | 0.0924 | 0.0105 | 0.0963 | 0.0110 |
| Co. Anxiety | 0.0416 | 0.0023 | 24543 | 249062 | 89366 | 0.0897 | 0.0825 | 0.85 | 0.55 | PMID: 11098417 | 0.1135 | 0.0063 | 0.1261 | 0.0070 | 0.1273 | 0.0072 |
| Non-co. anxiety | 0.0268 | 0.0022 | 16480 | 249062 | 61829 | 0.0621 | 0.0675 | 0.85 | 0.45 | | 0.0943 | 0.0077 | 0.1070 | 0.0088 | 0.1152 | 0.0096 |
| Early onset | 0.0489 | 0.0027 | 29292 | 250229 | 104890 | 0.1048 | 0.0563 | 0.85 | 0.375 | 3 first octiles | 0.0997 | 0.0055 | 0.1146 | 0.0063 | 0.1304 | 0.0071 |
| Late onset | 0.0153 | 0.0019 | 27796 | 250229 | 100068 | 0.1000 | 0.0563 | 0.85 | 0.375 | 3 last octiles | 0.0325 | 0.0040 | 0.0374 | 0.0046 | 0.0426 | 0.0053 |
| Recurrent | 0.0410 | 0.0025 | 30219 | 250229 | 107851 | 0.1078 | 0.0525 | 0.85 | 0.35 | PMID: | 0.0794 | 0.0048 | 0.0918 | 0.0056 | 0.1067 | 0.0066 |
| Single | 0.0222 | 0.0021 | 20973 | 250229 | 77404 | 0.0773 | 0.0975 | 0.85 | 0.65 | 20003092 | 0.0747 | 0.0071 | 0.0812 | 0.0077 | 0.0822 | 0.0083 |
| Suicidal | 0.0425 | 0.0025 | 40976 | 250229 | 140841 | 0.1407 | 0.0900 | 0.85 | 0.60 | PMID: | 0.0814 | 0.0048 | 0.0894 | 0.0053 | 0.0879 | 0.0053 |
| Non-suicidal | 0.0359 | 0.0023 | 37140 | 250229 | 129360 | 0.1292 | 0.0600 | 0.85 | 0.40 | 14628986 | 0.0625 | 0.0040 | 0.0715 | 0.0046 | 0.0798 | 0.0051 |
| Mild impair | 0.0222 | 0.0019 | 28721 | 250229 | 103055 | 0.1030 | 0.0300 | 0.85 | 0.20 | | 0.0372 | 0.0032 | 0.0439 | 0.0038 | 0.0600 | 0.0052 |
| Moderate impair | 0.0338 | 0.0025 | 28991 | 250229 | 103924 | 0.1038 | 0.0300 | 0.85 | 0.20 | PMID: 15930066 | 0.0562 | 0.0042 | 0.0665 | 0.0049 | 0.0908 | 0.0066 |
| Severe impair | 0.0382 | 0.0025 | 25825 | 250229 | 93636 | 0.0936 | 0.0900 | 0.85 | 0.60 | | 0.1042 | 0.0068 | 0.1145 | 0.0075 | 0.1127 | 0.0073 |
| Postpartum | 0.0250 | 0.0053 | 6333 | 95736 | 23760 | 0.0620 | 0.10 | 0.90 | 0.10 | PMID: 16260528 | 0.1131 | 0.0240 | 0.1131 | 0.0240 | 0.1073 | 0.0227 |
| CIDI-based MD | 0.0315 | 0.0024 | 17121 | 250229 | | 0.0640 | 0.15 | 0.85 | | | 0.1572 | 0.0120 | 0.1572 | 0.0120 | 0.1312 | 0.0102 |
| ICD-coded MD | 0.0350 | 0.0025 | 12135 | 250229 | | 0.0463 | 0.15 | 0.85 | | | 0.2370 | 0.0169 | 0.2370 | 0.0169 | 0.1983 | 0.0142 |
| Probable MD | 0.0299 | 0.0023 | 28628 | 250229 | | 0.1027 | 0.15 | 0.85 | | | 0.0970 | 0.0075 | 0.0970 | 0.0075 | 0.0811 | 0.0063 |
| Self-reported MD | 0.0628 | 0.0096 | 50857 | 250229 | | 0.1689 | 0.15 | 0.85 | | | 0.1338 | 0.0205 | 0.1338 | 0.0205 | 0.1069 | 0.0048 |
| Cardinal MD | 0.0579 | 0.0025 | 83774 | 250229 | | 0.2508 | 0.15 | 0.85 | | | 0.0921 | 0.0040 | 0.0921 | 0.0040 | 0.077 | 0.0034 |
| Broad MD | 0.0547 | 0.0024 | 124121 | 250229 | | 0.3316 | 0.15 | 0.85 | | | 0.0738 | 0.0032 | 0.0738 | 0.0032 | 0.0618 | 0.0027 |

[a]N_effective =4*p*(1-p)*N, with p = sample case proportion, N = total sample size
[b]Sample case prevalence = Ncase/(Ncase + Ncontrol)
[c]Population case prevalence = 0.15*case proportion (except PPD)
[d]Population control prevalence = 85% (except PPD). For MD phenotypes, population case prevalence = estimated lifetime prevalence of MD 15%, and population control prevalence = 85%
[e]Heritability and SE were converted from observed scale to liability scale using the method from Yap et al. (2018)[11]
[f]Heritability and SE were converted from observed scale to liability scale using the equation 17 in Lee at al. (2011)[10], same as used in LDSC software
[g]Heritability and SE on observed scale using based on 50:50 case-control ascertainment

### Table S7 Significant loci in MD subtypes and broad MD phenotype GWAS

| CHR | Region | rsID | POS | A1/A2 | AF1 | OR | SE | P | Nearest Gene | Mapped Gene(s) | Significant in previous large GWAS† | Significant in broad MD GWAS | P-values in GWAS of other subtype in comparison |
|---|---|---|---|---|---|---|---|---|---|---|---|---|---|
| **Non-atypical-like features** | | | | | | | | | | | | | **MD with atypical-like features** |
| 1 | 72511514-72838406 | rs66495454 | 72748567 | G/GTCCT | 0.62 | 0.9501 | 0.0073 | 1.82e-12 | NEGR1 | NEGR1 | PGC+ UKB, Top10K | Yes | 2.51e-03 |
| 6 | 27309779-29548089 | rs9393925 | 28682442 | T/C | 0.14 | 0.9424 | 0.0101 | 3.71e-09 | RPSAP2 | >10 genes | PGC+ UKB, Top10K | Yes | 1.18e-01 |
| 12 | 113349833-113349833 | rs55676265 | 113349833 | A/G | 0.79 | 0.9535 | 0.0086 | 3.40e-08 | OAS1:RP1-71H24.1 | OAS1 | | No | 7.98e-01 |
| 13 | 53608084-53842904 | rs2806950 | 53646197 | G/A | 0.38 | 1.0446 | 0.0073 | 1.80e-09 | OLFM4 | OLFM4 | Top10K | Yes | 6.54e-03 |
| 17 | 56424710-56451552 | rs56058331 | 56427128 | G/A | 0.58 | 0.9615 | 0.0071 | 3.12e-08 | BZRAP1-AS1:SUPT4H1 | SUPT4H1, RNF43 | * | No | 8.17e-01 |
| 19 | 4944555-5022857 | rs59562763 | 4969102 | G/GC | 0.65 | 0.9588 | 0.0074 | 1.04e-08 | KDM4B | KDM4B | * | Yes | 5.77e-02 |
| **Anxiety** | | | | | | | | | | | | | **Non-anxiety** |
| 1 | 72544704-72748567 | rs66495454 | 72748567 | G/GTCCT | 0.62 | 0.9465 | 0.0097 | 1.51e-08 | NEGR1 | NEGR1 | PGC+ UKB, Top10K | Yes | 5.36e-04 |
| 7 | 12233848-12286050 | rs3807866 | 12250378 | G/A | 0.59 | 0.9432 | 0.0095 | 7.67e-10 | TMEM106B | TMEM106B | PGC+ UKB, Top10K | Yes | 4.03e-03 |
| 9 | 11275587-11771159 | rs958538 | 11522951 | T/C | 0.75 | 1.0663 | 0.0109 | 3.72e-09 | RP11-23D5.1 | | PGC+ UKB, Top10K | No | 8.49e-01 |
| 14 | 103833065-104000802 | 14:103964545_TA_T | 103964545 | TA/T | 0.30 | 0.9443 | 0.0104 | 3.75e-08 | MARK3 | MARK3, CKB, TRMT61A | PGC+ UKB, Top10K | Yes | 1.73e-03 |
| **Early onset** | | | | | | | | | | | | | **Late onset** |
| 1 | 72511514-72838406 | rs7542974 | 72544704 | G/A | 0.75 | 0.9282 | 0.0100 | 7.79e-14 | NEGR1 | NEGR1 | PGC+ UKB, Top10K | Yes | 6.72e-02 |
| 2 | 34996836-35088235 | 2:35034680_CGA_C | 35034680 | CGA/C | 0.50 | 1.0496 | 0.0087 | 2.84e-08 | AC012593.1 | | * | No | 4.94e-02 |
| 3 | 85517112-86192846 | rs10865612 | 85868529 | T/C | 0.65 | 0.9380 | 0.0090 | 1.11e-12 | CADM2:CADM2-AS2 | CADM2 | * | No | 9.21e-01 |
| 6 | 27374680-27486633 | rs6914846 | 27410679 | A/G | 0.40 | 0.9522 | 0.0090 | 4.50e-08 | ZNF184 | ZNF391, ZNF184 | PGC+ UKB, Top10K | Yes | 8.07e-05 |
| 6 | 29530868-29530868 | rs362529 | 29530868 | C/T | 0.85 | 0.9347 | 0.0120 | 1.84e-08 | GABBR1 | UBD, GABBR1 | * | No | 2.41e-01 |
| 7 | 24548616-24801999 | rs2711093 | 24617290 | C/T | 0.70 | 0.9472 | 0.0095 | 1.00e-08 | MPP6 | MPP6, DFNA5 | | Yes | 2.67e-02 |
| 9 | 16713666-16784682 | rs35261422 | 16754554 | A/AT | 0.78 | 1.0607 | 0.0104 | 1.69e-08 | BNC2 | BNC2 | * | No | 9.07e-01 |

| | | | | | | | | | | | | | |
|---|---|---|---|---|---|---|---|---|---|---|---|---|---|
| 10 | 126711107-126738471 | rs34260682 | 126711499 | G/A | 0.91 | 1.0892 | 0.0156 | 4.51e-08 | CTBP2 | CTBP2 | | No | 6.83e-02 |
| 13 | 53608084-53675539 | rs3803259 | 53613331 | G/C | 0.48 | 1.0506 | 0.0087 | 1.72e-08 | OLFM4 | OLFM4 | Top10K | Yes | 2.25e-04 |
| 14 | 60179792-60663420 | rs216519 | 60653477 | C/A | 0.62 | 0.9518 | 0.0088 | 2.31e-08 | PSMA3P | RTN1, LRRC9, PCNXL4, DHRS7 | | No | 1.69e-01 |
| 16 | 13665985-13805809 | rs56225373 | 13755408 | T/G | 0.86 | 0.9264 | 0.0126 | 1.31e-09 | U95743.1 | | PGC+ UKB, Top10K | No | 4.79e-01 |
| 19 | 18182266-18279816 | rs11667720 | 18193751 | G/C | 0.57 | 0.9471 | 0.0090 | 1.52e-09 | IL12RB1 | IL12RB1, MAST3, PIK3R2, IFI30 | * | No | 6.28e-02 |
| 20 | 59827231-59827231 | rs143502921 | 59827231 | G/C | 0.82 | 0.9196 | 0.0126 | 2.48e-11 | CDH4 | CDH4 | * | Yes | 8.19e-02 |
| **Recurrent** | | | | | | | | | | | | | **Single** |
| 2 | 15311954-15468791 | rs6431690 | 15443525 | T/C | 0.55 | 1.0530 | 0.0086 | 1.76e-09 | NBAS | NBAS | | No | 3.22e-04 |
| 2 | 152886708-153120132 | rs62175074 | 152886708 | T/C | 0.79 | 0.9415 | 0.0109 | 3.31e-08 | CACNB4 | CACNB4 | * | No | 2.41e-02 |
| 2 | 212702426-212778384 | rs74338595 | 212749786 | T/C | 0.71 | 1.0534 | 0.0094 | 2.87e-08 | ERBB4 | ERBB4 | | No | 8.71e-01 |
| 5 | 164467717-164588817 | rs543533963 | 164474868 | C/T | 0.45 | 0.9533 | 0.0087 | 3.98e-08 | CTC-340A15.2 | | PGC+ UKB, Top10K | No | 2.81e-01 |
| 6 | 26309908-29607101 | rs1233393 | 29549653 | C/G | 0.87 | 1.0869 | 0.0127 | 5.98e-11 | GABBR1 | >10 genes | PGC+ UKB, Top10K | Yes | 2.45e-03 |
| 11 | 88472034-88978545 | rs1942496 | 88840925 | G/A | 0.51 | 1.0513 | 0.0086 | 5.81e-09 | AP001482.1 | GRM5, TYR | PGC+ UKB, Top10K | Yes | 3.86e-03 |
| 19 | 31891006-31927547 | rs2111530 | 31891006 | A/G | 0.61 | 0.9513 | 0.0088 | 1.20e-08 | AC007796.1 | | | Yes | 4.59e-01 |
| **Suicidal** | | | | | | | | | | | | | **Non-suicide** |
| 1 | 109873290-110040460 | rs11590351 | 110035159 | T/C | 0.75 | 0.9516 | 0.0086 | 8.54e-09 | ATXN7L2 | SORT1, PSMA5, SYPL2, ATXN7L2, CYB561D1, AMIGO1 | | No | 4.77e-03 |
| 4 | 2412967-2439670 | rs113065538 | 2420428 | C/A | 0.35 | 1.0441 | 0.0078 | 3.41e-08 | ZFYVE28 | ZFYVE28 | | No | 2.33e-02 |
| 11 | 90528418-90646073 | rs10830592 | 90592959 | A/G | 0.32 | 0.9565 | 0.0080 | 2.43e-08 | DISC1FP1 | | | No | 6.31e-02 |
| **Non-suicidal** | | | | | | | | | | | | | **Suicide** |
| 1 | 72544704-72838406 | rs66495454 | 72748567 | G/GTCCT | 0.62 | 0.9510 | 0.0081 | 5.79e-10 | NEGR1 | NEGR1 | PGC+ UKB, Top10K | Yes | 8.62e-08 |
| 1 | 239697408-239760514 | rs12118109 | 239760514 | G/A | 0.96 | 0.8982 | 0.0197 | 4.68e-08 | CHRM3 | CHRM3 | | No | 8.03e-01 |
| 6 | 27374680-27486633 | rs6914846 | 27410679 | A/G | 0.40 | 0.9559 | 0.0081 | 2.96e-08 | ZNF184 | ZNF391, ZNF184 | PGC+ UKB, Top10K | Yes | 2.18e-05 |

| | | | | | | | | | | | | | | |
|---|---|---|---|---|---|---|---|---|---|---|---|---|---|---|
| 10 | 76463067-76506933 | rs9733673 | 76504584 | G/T | 0.85 | 0.9358 | 0.0113 | 4.68e-09 | RP11-383B24.3 | ADK | | No | 9.58e-01 | |
| 15 | 83170194-83337059 | rs35385880 | 83214764 | C/CT | 0.35 | 0.9545 | 0.0082 | 1.27e-08 | RP11-152F13.10:RP11-379H8.1:CPEB | RPS17L, RP11-152F13.10, RP11-379H8.1, CPEB1, AP3B2 | * | No | 1.34e-03 | |
| **Moderate impairment** | | | | | | | | | | | | | **Mild** | **Severe** |
| 1 | 72748567-72838406 | rs66495454 | 72748567 | G/GTCCT | 0.62 | 0.9428 | 0.0090 | 6.95e-11 | NEGR1 | NEGR1 | PGC+ UKB, Top10K | Yes | 4.23e-03 | 6.81e-09 |
| 1 | 243197475-243500994 | rs4658548 | 243394093 | C/T | 0.63 | 1.0520 | 0.0090 | 1.82e-08 | CEP170:AC092782.1 | CEP170, AC092782.1, SDCCAG8 | | No | 1.25e-02 | 5.54e-02 |
| 16 | 10472145-10596428 | rs141035531 | 10589515 | C/G | 0.98 | 0.8377 | 0.0311 | 1.28e-08 | RNU6-633P | ATF7IP2 | * | No | 2.25e-03 | 2.45e-01 |
| **Severe impairment** | | | | | | | | | | | | | **Mild** | **Moderate** |
| 1 | 72511514-72748567 | rs7542974 | 72544704 | G/A | 0.75 | 0.9402 | 0.0106 | 6.37e-09 | NEGR1 | NEGR1 | PGC+ UKB, Top10K | Yes | 1.21e-03 | 2.18e-06 |
| 5 | 164334125-164678946 | rs10476484 | 164438264 | A/G | 0.73 | 0.9399 | 0.0103 | 2.13e-09 | CTC-340A15.2 | | PGC+ UKB, Top10K | No | 4.73e-01 | 1.73e-01 |
| 6 | 27321848-27476592 | rs6456788 | 27422066 | C/T | 0.35 | 0.9485 | 0.0096 | 3.32e-08 | ZNF184 | ZNF391, ZNF184 | PGC+ UKB, Top10K | Yes | 1.82e-03 | 3.48e-04 |
| 6 | 27868792-29548089 | rs9393925 | 28682442 | T/C | 0.14 | 0.9290 | 0.0131 | 2.01e-08 | RPSAP2 | >10 genes | PGC+ UKB, Top10K | Yes | 4.04e-03 | 1.53e-02 |
| 13 | 53608084-53675691 | rs12552 | 53625781 | A/G | 0.44 | 1.0603 | 0.0093 | 2.55e-10 | OLFM4 | | Top10K | Yes | 1.21e-01 | 2.16e-07 |
| **PPD** | | | | | | | | | | | | | | |
| 2 | 189234143-189682431 | rs11683671 | 189669039 | C/A | 0.93 | 0.8152 | 0.0367 | 2.68e-08 | AC079613.1 | GULP1 | | No | - | |
| **MD broad phenotype** | | | | | | | | | | | | | | |
| 1 | 37663818-37680479 | rs215849 | 37677592 | C/G | 0.76 | 0.9669 | 0.0058 | 5.95e-09 | RNU6-636P | | * | - | - | |
| 1 | 71815804-73699761 | rs66495454 | 72748567 | G/GTCCT | 0.62 | 0.9593 | 0.0050 | 9.32e-17 | NEGR1 | NEGR1 | PGC+ UKB, Top10K | - | - | |
| 1 | 197342380-197813558 | rs12123169 | 197780966 | T/A | 0.79 | 1.0360 | 0.0059 | 2.51e-09 | DENND1B | CRB1, DENND1B | PGC+ UKB, Top10K | - | - | |
| 2 | 23178683-23183651 | rs6744716 | 23181209 | T/A | 0.69 | 0.9716 | 0.0052 | 3.02e-08 | AC016768.1 | | | - | - | |
| 2 | 34996836-35092802 | rs2083210 | 35027554 | G/T | 0.56 | 1.0275 | 0.0049 | 2.62e-08 | AC012593.1 | | | - | - | |
| 2 | 155654466-156116188 | rs1037094 | 155656548 | C/A | 0.79 | 1.0338 | 0.0060 | 2.50e-08 | KCNJ3 | KCNJ3 | | - | - | |
| 5 | 87981030-88175251 | rs28712135 | 88084016 | T/A | 0.53 | 1.0288 | 0.0049 | 6.33e-09 | MEF2C | MEF2C | Top10K* | - | - | |

| | | | | | | | | | | | | | |
|---|---|---|---|---|---|---|---|---|---|---|---|---|---|
| 5 | 103671867-104082179 | rs30266 | 103972357 | G/A | 0.67 | 0.9699 | 0.0051 | 2.92e-09 | RP11-6N13.1 | | PGC+ UKB, Top10K | - | - |
| 6 | 25417423-29607101 | rs10807023 | 27410220 | G/A | 0.38 | 0.9644 | 0.0050 | 2.79e-13 | ZNF184 | >10 genes | PGC+ UKB, Top10K | - | - |
| 7 | 12233848-12286050 | rs3807865 | 12250402 | G/A | 0.59 | 0.9731 | 0.0049 | 2.46e-08 | TMEM106B | TMEM106B | PGC+ UKB, Top10K | - | - |
| 7 | 24590979-24779987 | rs2529047 | 24598708 | G/A | 0.70 | 0.9696 | 0.0053 | 6.20e-09 | MPP6 | MPP6, DFNA5 | | - | - |
| 11 | 31808280-31856654 | rs7942007 | 31835418 | A/G | 0.81 | 0.9663 | 0.0062 | 3.43e-08 | PAX6:RCN1 | ELP4, PAX6, RCN1 | Top10K | - | - |
| 11 | 88470041-89026407 | 11:88769807 | 88769807 | GGCAAG ACTAT TAATCAG TATTT GTACTCA AATTT GCCCCAA AGTC CCTCAAC TA/G | 0.42 | 1.0317 | 0.0049 | 2.52e-10 | GRM5 | GRM5, TYR | PGC+ UKB, Top10K | - | - |
| 13 | 53608084-53675539 | rs3803259 | 53613331 | G/C | 0.48 | 1.0327 | 0.0049 | 5.84e-11 | OLFM4 | OLFM4 | PGC+ UKB, Top10K | - | - |
| 13 | 69779070-70024608 | rs7989827 | 69857608 | T/G | 0.52 | 1.0273 | 0.0048 | 2.47e-08 | LINC00383 | | | - | - |
| 14 | 60155307-60247284 | 14:60156702 _CT_C | 60156702 | CT/C | 0.35 | 1.0294 | 0.0052 | 2.27e-08 | RTN1 | RTN1 | * | - | - |
| 14 | 103833065-104093521 | rs2071408 | 103987078 | G/A | 0.63 | 1.0333 | 0.0050 | 7.46e-11 | CKB | MARK3, CKB, TRMT61A, BAG5, KLC1, RP11-73M18.2, APOPT1 | PGC+ UKB, Top10K | - | - |
| 15 | 74010301-74062710 | rs4327001 | 74010301 | G/A | 0.59 | 0.9732 | 0.0049 | 3.24e-08 | CD276 | CD276, C15orf59 | | - | - |
| 18 | 50364022-51059160 | rs112633410 | 50740013 | G/A | 0.57 | 0.9713 | 0.0049 | 2.51e-09 | DCC | DCC | PGC+ UKB, Top10K | - | - |
| 19 | 4944555-5022857 | rs58324296 | 4969199 | A/C | 0.65 | 0.9715 | 0.0051 | 1.18e-08 | KDM4B | KDM4B | | - | - |
| 19 | 31891006-31927547 | rs2111530 | 31891006 | A/G | 0.60 | 0.9732 | 0.0050 | 4.25e-08 | AC007796.1 | | | - | - |
| 20 | 59827231-59827231 | rs143502921 | 59827231 | G/C | 0.82 | 0.9539 | 0.0070 | 2.08e-11 | CDH4 | CDH4 | * | - | - |

*Some variants in those loci were not analyzed in PGC+UKB GWAS, which might be excluded in quality control or not examined.

†Top10K: Data contains genome-wide summary statistics of the most informative 10,000 variants from a meta-analysis of the 33 cohorts of the Psychiatric Genomics Consortium, the broad depression phenotype in the full release of the UK Biobank and 23andMe data

PGC+UKB: Data contains genome-wide summary statistics from a meta-analysis of the 33 cohorts of the Psychiatric Genomics Consortium (excluding UK Biobank and 23andMe data) and the broad depression phenotype in the full release of the UK Biobank

Citation: Howard et al (2019) [dataset][27].

Col 1 (CHR): Chromosome of the locus

Col 2 (Region): Start and end position of the locus

Col 3 (rsID): RsID of the lead SNP

Col 4 (POS): Position of lead SNP on hg19

Col 5 (A1/A2): Effect/non-effect allele of the lead SNP

Col 6 (AF1): Allele frequency of effect allele of the lead SNP in study sample

Col 7 (OR): Odd ratio of the lead SNP which was converted from beta coefficients in the GWAS result after correcting for sample case-control ascertainment

Col 8 (SE): Standard error of log(OR)

Col 9 (P): P-value of the lead SNP

Col 10 (Nearest Gene): The nearest Gene of the lead SNP based on ANNOVAR annotations

Col 11 (Mapped gene): Positional mapped gene. Loci was mapped to genes up to 10 kb

Col 12 (Significant in previous large GWAS): If the SNPs were significant in the previous 2 large GWAS, Top10K and PGC+UKB

Col 13 (Significant in broad MD GWAS): Checked if the subtype-specific significant SNPs were also significant in the broad MD GWAS

Col 14 (P-values in GWAS of other subtype in comparison): P-values of the SNPs in the GWAS of the other subtype in the same comparison group

### Table S8 SNP-heritability and genetic correlation for subtypes within CIDI-SF MD cases

| | N case[a] | N control[b] | Sample case prevalence[c] | N effective | $h^2_{SNP}$ observed scale[d] | SE observed scale[d] | $h^2_{SNP}$ observed 50/50[e] | *SE* observed 50/50[e] | $h^2_{SNP}$ liability Yap[f] | SE liability Yap[f] | $r_g$ | Se $r_g$ | 95% CI $r_g$ |
|---|---|---|---|---|---|---|---|---|---|---|---|---|---|
| Atypical-like features CIDI | 1325 | 250229 | 0.0053 | 5272 | 0.0066 | 0.0019 | 0.3172 | 0.0884 | 0.2209 | 0.0636 | 0.5912 | 0.0942 | 0.4066; |
| Non-atypical-like features CIDI | 11928 | 250229 | 0.0455 | 45541 | 0.0215 | 0.0022 | 0.1236 | 0.0129 | 0.123 | 0.0126 | | | 0.7758 |
| Severe CIDI | 1175 | 250229 | 0.0047 | 4678 | 0.0010 | 0.0022 | 0.0537 | 0.1162 | 0.0457 | 0.1006 | 0.9781 | 0.2724 | 0.4442; |
| Mild/moderate CIDI | 1424 | 250229 | 0.0057 | 5664 | 0.0020 | 0.0016 | 0.0885 | 0.0692 | 0.0755 | 0.0604 | | | 1.5120 |
| Co. anxiety CIDI | 7303 | 249062 | 0.0285 | 28380 | 0.0217 | 0.0023 | 0.1965 | 0.021 | 0.1746 | 0.0185 | 0.7739 | 0.0634 | 0.6496; |
| Non-co. Anxiety CIDI | 5920 | 249062 | 0.0232 | 23130 | 0.0125 | 0.002 | 0.1383 | 0.0217 | 0.1131 | 0.0181 | | | 0.8982 |
| Early CIDI | 6570 | 250229 | 0.0256 | 25608 | 0.0210 | 0.0022 | 0.2104 | 0.0225 | 0.161 | 0.0169 | 0.6021 | 0.084 | 0.4375; |
| Late CICI | 5498 | 250229 | 0.0215 | 21519 | 0.0066 | 0.0018 | 0.0788 | 0.0213 | 0.0600 | 0.0164 | | | 0.7667 |
| Recurrent CIDI | 11094 | 250229 | 0.0425 | 42492 | 0.0265 | 0.0023 | 0.1632 | 0.0139 | 0.1213 | 0.0105 | 0.8402 | 0.0947 | 0.6546; |
| Single CIDI | 5602 | 250229 | 0.0219 | 21917 | 0.0091 | 0.0018 | 0.1057 | 0.021 | 0.102 | 0.0202 | | | 1.0258 |
| Suicide CIDI | 10492 | 250229 | 0.0402 | 40279 | 0.0226 | 0.0022 | 0.1464 | 0.014 | 0.1356 | 0.0132 | 0.7641 | 0.0802 | 0.6069; |
| Non-suicide CIDI | 5500 | 250229 | 0.0215 | 21527 | 0.0118 | 0.002 | 0.1404 | 0.0234 | 0.1098 | 0.0186 | | | 0.9213 |
| PPD CIDI | 1588 | 95736 | 0.0163 | 6248 | 0.0131 | 0.0046 | 0.2046 | 0.0712 | 0.2149 | 0.0754 | | | |

[a]N case = number of cases as defined in *Supplementary table S4* and was used in GWAS

[b]N control = number of controls as defined in *Supplementary table S2* and was used in GWAS

[c]Sample case prevalence = N case/(N case + N control)

[d]$h^2_{SNP}$ and standard error (SE) were estimated on observed scale

[e] $h^2_{SNP}$ and SE were estimated on observed scale based on 50:50 case-control ascertainment

[f]$h^2_{SNP}$ and SE were converted from observed (see [d]) to liability scale using the same population case and control as in *Supplementary table S6* using the conversion method in Yap et al. (2018)[11]

### Table S9 Compare subtype $h^2$ between primary analysis and independent controls analysis

| Subtype | Primary analysis $h^2_{SNP}$ observed scale[a] (se) | Primary analysis $h^2_{SNP}$ liability Yap[b] (CI) | Primary analysis $h^2_{SNP}$ observed 50/50[c] (CI) | Independent control analysis $h^2_{SNP}$ observed scale[a] (se) | Independent control analysis $h^2_{SNP}$ liability Yap[b] (CI) | Independent control analysis P-value[d] | Independent control analysis $h^2_{SNP}$ observed 50/50[c] (CI) | Independent control analysis P-value[d] |
|---|---|---|---|---|---|---|---|---|
| Early onset | 0.049 (0.003) | 0.100 (0.089-0.110) | 0.130 (0.116-0.144) | 0.078 (0.005) | 0.097 (0.086-0.108) | 2.306e-19 | 0.127 (0.113-0.142) | 1.400e-19 |
| Late onset | 0.015 (0.002) | 0.033 (0.025-0.040) | 0.043 (0.032-0.053) | 0.025 (0.004) | 0.033 (0.024-0.042) | | 0.043 (0.031-0.054) | |
| Suicidal | 0.043 (0.003) | 0.081 (0.072-0.091) | 0.088 (0.078-0.098) | 0.063 (0.004) | 0.078 (0.068-0.088) | 0.017 | 0.085 (0.074-0.095) | 0.520 |
| Non-suicidal | 0.036 (0.002) | 0.063 (0.055-0.070) | 0.080 (0.070-0.090) | 0.056 (0.004) | 0.062 (0.054-0.071) | | 0.080 (0.069-0.090) | |
| Severe impaired | 0.038 (0.003) | 0.104 (0.091-0.118) | 0.113 (0.098-0.127) | 0.081 (0.006) | 0.103 (0.089-0.118) | Severe-moderate 0.002e-09 | 0.112 (0.097-0.128) | Severe-moderate 0.017 |
| Moderate impaired | 0.034 (0.003) | 0.056 (0.048-0.064) | 0.091 (0.078-0.104) | 0.067 (0.005) | 0.054 (0.046-0.062) | Severe-mild 7.633e-15 | 0.087 (0.074-0.101) | Severe-mild 1.245e-06 |
| Mild impaired | 0.022 (0.002) | 0.037 (0.031-0.043) | 0.060 (0.050-0.070) | 0.048 (0.005) | 0.039 (0.031-0.047) | Moderate-mild 0.009 | 0.063 (0.050-0.075) | Moderate-mild 0.008 |

*[a]SNP-$h^2$ on observed scale*

*[b]SNP-$h^2$ on liability scale, converted from observed scale using method in Yap et al. (2018)[11]*

*[c]SNP-$h^2$ on observed scale based on 50:50 case-control ascertainment*

*[d]P-value derived from the Delta method which tests significant difference between estimates on liability scale and observed-$h^2$ 50/50*

### III. SUPPLEMENTARY FIGURES

**Figure S1 Included sample of study**

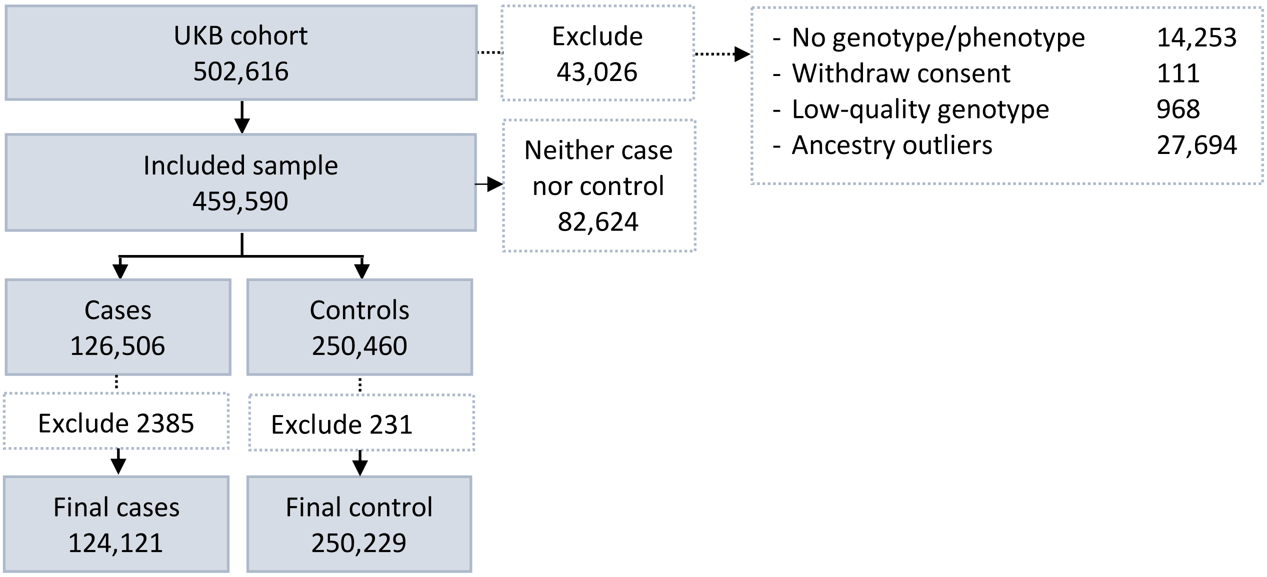

***Figure S1*** *Included sample*

The dataset from UKB contains 502 616 observations. We excluded those who 1) did not have both genotype and phenotype data. 2) withdraw consent, 3) Low-quality genotype data, and 4) European ancestry outliers. Individuals without European ancestry were excluded by removing those whose first two principal components exceeded three standard deviations from the means of 1000 Genome European samples[28]. We retained 459 590 observations.

We defined MD cases and controls as in *Supplementary table S2*. We identified 126,506 individuals who met at least one of the five MD case criteria. We used seven exclusion criteria to define controls (N=250 460) including five case criteria, and two additional criteria of help seeking for MD (N=38 177) and using antidepressant (N=175 367). Individuals who met the criteria for help seeking MD and using antidepressants thus fell into the 'neither case nor control' group.

**Figure S2 SNP- heritability for MD definitions in UKB**

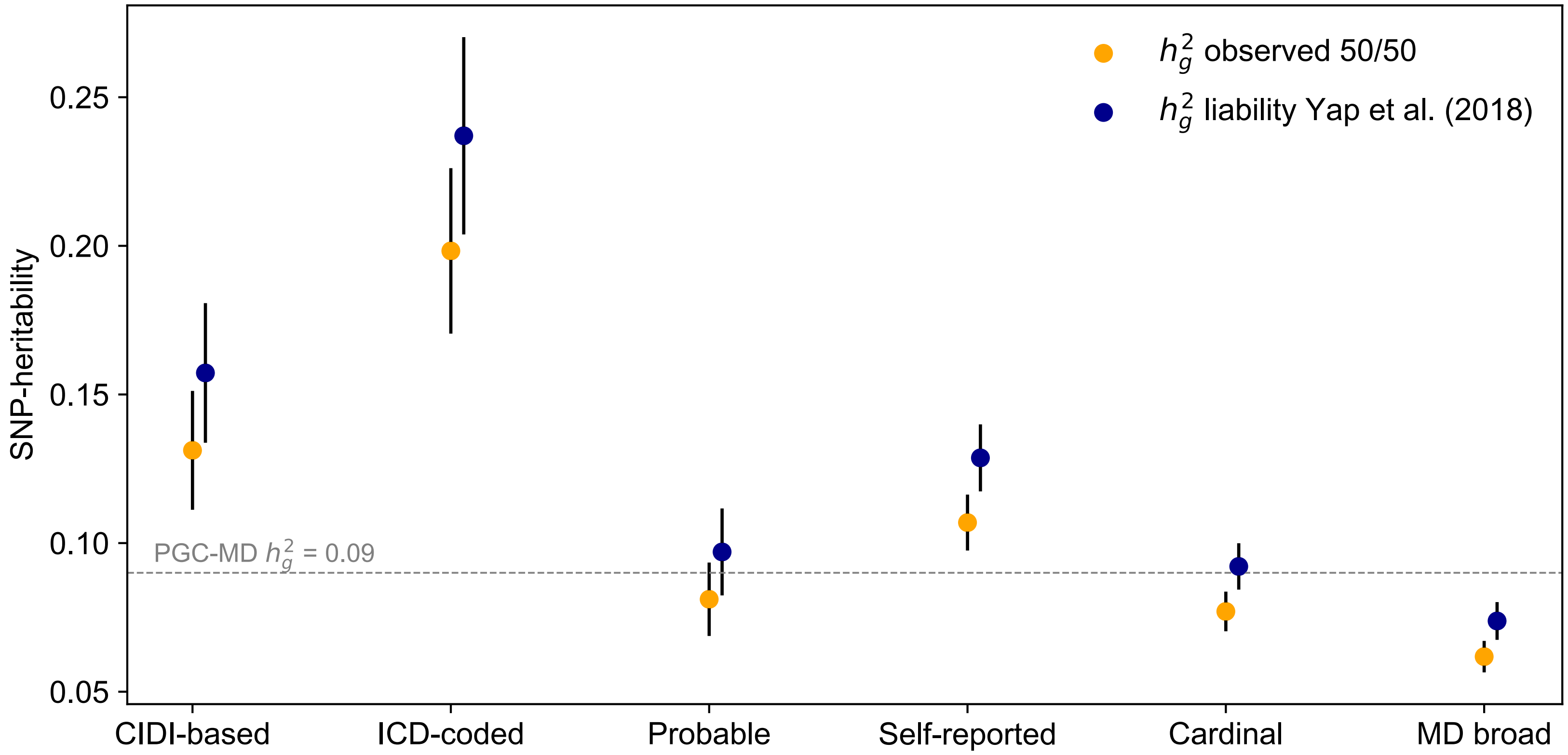

**Figure S2** SNP-heritability for MD definitions in UKB
Orange markers show $h^2_{SNP}$ estimates on observed scale based on 50:50 case-control ascertainment[9]. **Blue** markers show point $h^2_{SNP}$ estimates on liability scale, converted from observed scale using formula in Yap et al. (2018)[11]. Error bars show 95% confidence intervals.

**Figure S3 SNP-heritability on liability scale using different conversion methods**

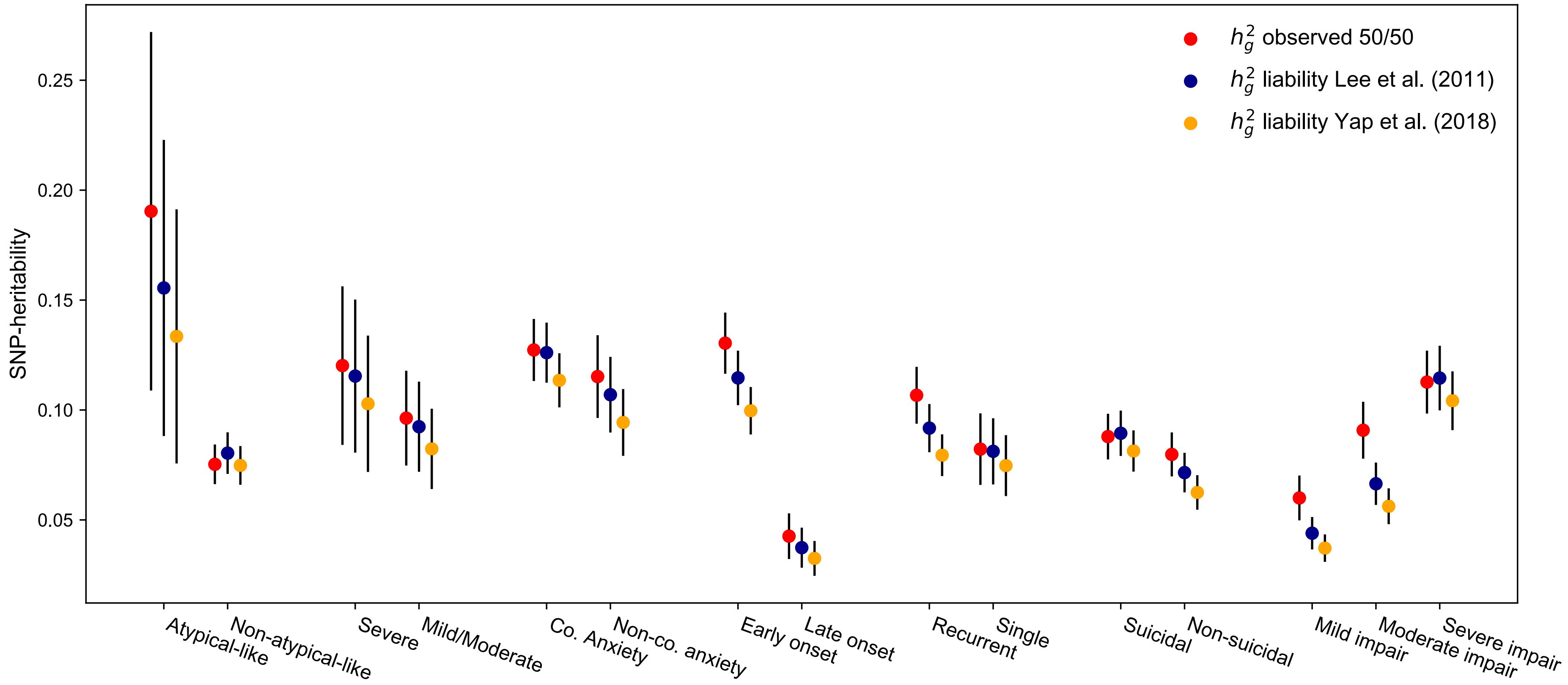

**Figure S3**: SNP-heritability on liability scale using different conversion methods
The point presents *SNP-h*$^2$. Error bars show 95% CI. **Red**: *SNP-h*$^2$ on observed scale based on 50:50 case-control ascertainment. **Blue**: *SNP-h*$^2$ transformed from observed to liability scale using the standard conversion from Lee et al. (2011)[10]. **Orange**: *SNP-h*$^2$ transformed from observed to liability scale based on the conversion from Yap et al. (2018)[11] which takes into account extreme phenotype selection.

**Figure S4 Subtype SNP-heritability estimated among all samples and unrelated samples**

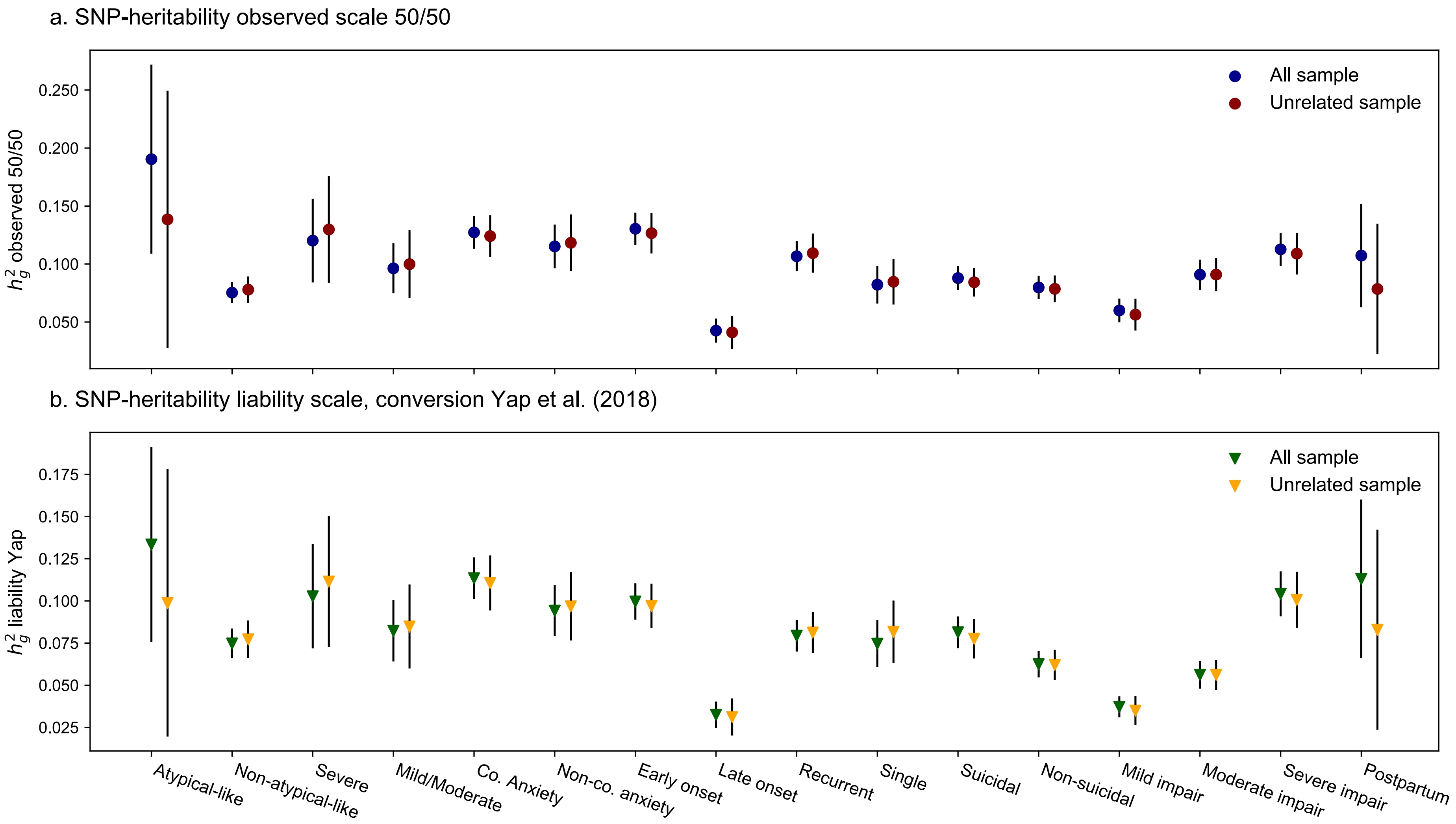

**Figure S4**: Subtype SNP-heritability estimated among all sample (GWAS using fastGWA) and unrelated sample (GWAS using PLINK). a. Estimates on observed scale based on 50:50 case-control ascertainment. b. Estimates on liability scale, converted from observed to liability using method from Yap et. al (2018)[11]. Error bars show 95% CI of the estimates.

### Figure S5 Phenotypic and genetic correlation across MD subtypes

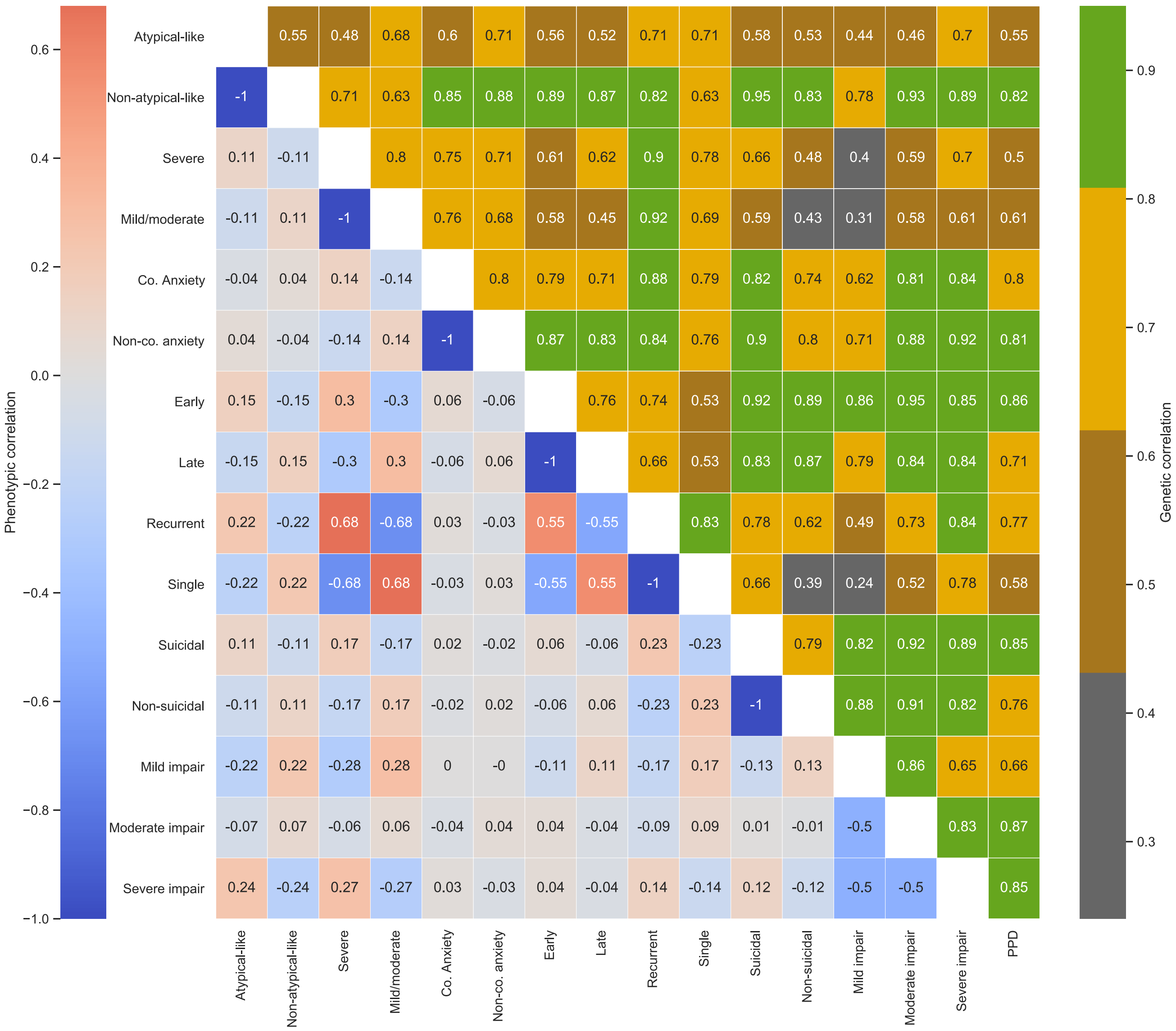

**Figure S5**: Phenotypic and genetic correlation across MD subtypes. **Upper triangle**: pair-wise genetic correlations. **Lower triangle**: Phenotypic correlation for subtype cases only (because of common control group). "-1" indicates that subtype definitions were mutually exclusive (for subtype within same comparison group).
